# Disruption of seasonal influenza circulation and evolution during two pandemic events in Southeastern Asia: contrasting the role of human movement and pathogen competition

**DOI:** 10.1101/2024.06.19.24309151

**Authors:** Zhiyuan Chen, Joseph L.-H. Tsui, Jun Cai, Cécile Viboud, Louis du Plessis, Philippe Lemey, Moritz U. G. Kraemer, Hongjie Yu

**Affiliations:** School of Public Health, Key Laboratory of Public Health Safety, Ministry of Education, Fudan University, Shanghai, China; Department of Biology, University of Oxford, Oxford, UK; Fogarty International Center, National Institutes of Health, Bethesda, MD, USA; Department of Biosystems Science and Engineering, ETH Zürich, Basel, Switzerland; Swiss Institute of Bioinformatics, Lausanne, Switzerland; Department of Microbiology, Immunology and Transplantation, Rega Institute, KU Leuven, Leuven, Belgium; Pandemic Sciences Institute, University of Oxford, Oxford, UK

**Keywords:** Seasonal influenza, pandemics, phylogeography, circulation network, human movements, subtype interactions

## Abstract

East, South, and Southeast Asia (together referred to as Southeastern Asia hereafter) have been recognized as critical areas fuelling the global circulation of seasonal influenza. However, the internal migration network of seasonal influenza within Southeastern Asia remains unclear, including how pandemic-related disruptions altered the network structure and circulation dynamics in this region. Here, we leveraged genetic, epidemiological, and airline travel data between 2007-2023 to characterise the multiyear dispersal patterns of influenza A/H3N2 and B/Victoria viruses both out of and within Southeastern Asia, including during seasons marked by perturbations such as the 2009 A/H1N1 and COVID-19 pandemics. We show consistent Autumn-Winter movement waves of A/H3N2 and B/Victoria from Southeastern Asia to temperate regions during interpandemic seasons. During the COVID-19 pandemic this trend was interrupted for both subtypes, however the A/H1N1 pandemic only disrupted A/H3N2 spread. For influenza strains circulating in Southeastern Asia, we find a higher persistence of A/H3N2 than B/Victoria. We find pandemic-related disruptions in A/H3N2 antigenic evolution, with a greater time-advanced antigenic evolution during the 2009 A/H1N1 pandemic, and a greater time-lagged pattern during the COVID-19 pandemic, compared to inter-pandemic levels. Internally, in comparison to the interpandemic seasons, the inferred dispersal rates within Southeastern Asia decreased by 54.7% and 79.2% during the 2009 A/H1N1 and COVID-19 pandemic seasons, respectively; further, the internal movement structure within Southeastern Asia markedly diverged during the COVID-19 pandemic season, and to a lesser extent, during the 2009 A/H1N1 pandemic season. Analyses of the trunk location and phylogenetic similarity further reveal a temporally varying pattern within Southeastern Asia, suggesting a complex source-sink network, with a notable decrease in the mixing of lineages around the COVID-19 pandemic season. Our findings provide insights into the heterogeneous interplay between influenza circulation in Southeastern Asia and two distinct pandemic-related disruptions (strong decline in human movements during the COVID-19 pandemic, pronounced pathogen interference during the A/H1N1 pandemic), which can help anticipate the effects of potential mitigation strategies and the emergence of future influenza pandemic strains on influenza dynamics.

## Main text

Seasonal influenza infections occur annually and cause a significant disease burden across the world^1^. Human mobility and interconnectedness is thought to be the main driver of worldwide human influenza virus spread^2^, while a combination of antigenic evolution to escape immunity and waning immunity results in an oscillating supply of susceptible hosts leading to frequent reinfections^3^. Seasonal influenza viruses cause predictable annual epidemics in temperate regions as well as relatively divergent waves in tropical regions^4,5^. Newly-emerged influenza viruses can disrupt this pattern through cross-subtype population immunity^6,7^. In parallel, other co-circulating seasonal or novel respiratory pathogens can shape the spread of seasonal influenza viruses, especially when they are associated with human behavioural changes in response to non-pharmaceutical interventions (NPIs) and declarations of public health emergencies^8,9^. Two emerging pathogens, the swine-origin influenza A/H1N1 virus and SARS-CoV-2, triggered global pandemics in 2009 and 2020 respectively. However, whether and how changes in human behaviour and cross-subtype population immunity during the course of these pandemics affected seasonal influenza circulation, especially in Southeastern Asia (here defined as East, South, and Southeast Asia), remains unclear.

Our previous work evaluated the global dispersal patterns of seasonal influenza among 12 broad geographical regions prior, during, and after the COVID-19 pandemic, and found that the pandemic’s onset led to a shift in the intensity and structure of the international movement of influenza lineages^10^. Beyond the global perspective focusing only on the COVID-19 or A/H1N1 pandemics alone^7,8,10^, comparisons of the impacts of the 2009 A/H1N1 and COVID-19 pandemics on influenza circulation at a finer spatial scale are still lacking. Apart from the heterogeneity in the intensity of NPIs during the two pandemics, different degrees of viral interference (namely, virus‒virus interactions via cross-immunity) can also be expected between seasonal influenza viruses and the two pandemic viruses^6,11,12^. Additionally, the structure of the influenza virus migration network within Asia, especially Southeastern Asia, has rarely been explored. Understanding the internal migration network within Southeastern Asia is critical as it has been suggested to play an essential role in generating antigenically distinct seasonal viruses and seeding global seasonal influenza epidemics^4,13,14^. Changes in human behaviour and cross-immunity during the 2009 A/H1N1 and COVID-19 pandemics provide natural experiments to evaluate temporal shifts on the circulation patterns of seasonal influenza in Southeastern Asia relative to the baseline interpandemic period, and elucidate the mechanisms at play. A comprehensive genomic and epidemiological assessment of the interplay between pandemic-related disruptions and seasonal influenza circulation in Southeastern Asia can further inform potential strategies for mitigating global disease burdens in the future.

In this study, we leveraged genetic, epidemiological, and airline travel data to assess the circulation dynamic of seasonal influenza emanating from and within Southeastern Asia between 2007 and 2023, covering the apexes of two global pandemics and multiple interpandemic seasons. Specifically, we first inferred the seasonality of viral movement out of Southeastern Asia, and identified those viral lineages that potentially persisted within Southeastern Asia (defined as persistent lineages) to trace their internal circulation dynamics. We subsequently performed long-term comparisons of the internal spread of seasonal influenza within Southeastern Asia and evaluated how influenza circulation was impacted by the two pandemics.

## Methods

To develop our methods for epidemiological and genomic assessment, we combined epidemiological data, genetic data and airline data, together with a phylodynamic framework to infer viral movements and the emergence of antigenic novelty at various spatial levels.

### Epidemiological data

Global virological surveillance data for seasonal influenza was retrieved from FluNet, based on the WHO-led Global Influenza Surveillance and Response System (GISRS)^15^ and collated according to the methodology followed in^10^. We extracted the weekly number of specimens processed for influenza testing and positive detections by subtypes or lineages to calculate weekly positivity rates. We defined the influenza season in the southern hemisphere as running from ISO week 1 to week 52 of one year, whereas the influenza season in other regions was defined as running from ISO week 27 of one year to ISO week 26 of the next year. In each region, we defined an average seasonal pattern for inter-pandemic seasons based on the total positivity rates *p_ij_* of each seasonal influenza virus subtype/lineage in week *i* during influenza season *j*. We averaged the positivity rates *p_ij_* across interpandemic seasons after aligning curves based on the median week of peak occurrence with outlier seasons removed (for details see **Supplemental Figs. 1-2**)^16,17^. Since Southeastern Asia experienced two A/H3N2 waves during interpandemic seasons we performed two epidemic alignments per season by splitting each influenza season into a summer (ISO week 14 to 39) and winter season (ISO week 40 to ISO week 13 of the next year).

### Collation and sub-sampling of viral sequence data

We focused on A/H3N2 and B/Victoria in this study, because A/H1N1pdm09 only emerged in 2009 and therefore cannot be regarded as a seasonal influenza virus during the 2009 pandemic season^18^, and B/Yamagata potentially disappeared after March 2020^19^. A global genetic dataset of seasonal influenza sequences (hemagglutinin (HA) segment) sampled between 2007 and 2023 was retrieved from GISAID and GenBank on 17 Jan 2024, with details of data processing in^10^. To retain more sequences than our previous study^10^, we also included sequences with incomplete collection dates and only discarded poor quality sequences, as defined by the quality control criteria in Nextclade^20,21^.

Geographic locations of sequences were classified into Southeastern Asia and temperate regions, according to previous understanding of the global influenza circulation network^4^. Based on previous work on transmission characteristics of influenza and availability of genetic data in Asia^22,23^, here we defined the geographic scale of “Southeastern Asia” as comprising the entirety of Southeast Asia, parts of East Asia (China, Japan, South Korea), and parts of South Asia (Bangladesh, Bhutan, India, Sri Lanka, Nepal), which is in line with one of the seven influenza transmission zones defined in a previous clustering analysis^22^.

To trace the internal transmission network of influenza within Southeastern Asia, we used finer-grained internal spatial demes (**Extended Data Fig. 1**), where the majority of demes are set at country levels. Further, we divided mainland China into three zones, based on previous work showing heterogeneous seasonality patterns driven by climatic conditions (winter peak in north China, semi-annual peaks in central China, and spring peak in south China)^24,25^. In addition, in China, we separated Hong Kong, Macao, and Taiwan due to their distinct positions in the global air transport network^26^ and heterogeneous non-pharmaceutical interventions (NPIs) adopted during both pandemic interruptions compared to mainland China^27^. Finally, we combined Indonesia and East Timor, as well as Malaysia and Brunei, due to the low availability of publicly available genetic data in these neighbouring countries. This resulted in a total of 22 “country/sub-location level” (referred to as sub-location level hereafter) demes within Southeastern Asia.

Temperate regions comprised five sub-regions: North America (Canada and USA only) and Europe (Russia excluded) in the northern hemisphere (NH); Oceania (Australia and New Zealand only), South America, and the Southern part of Africa in the southern hemisphere (SH) (**Extended Data Fig. 1**). The entirety of South America was classified as a Southern Hemisphere temperate zone, given the recommended use of the Southern Hemisphere vaccine formulation throughout the region^28^ and a similar seasonality pattern across the whole region^5^. We recognize that tropical land masses exist in the northern part of South America.

To reduce the impact of sampling biases while maintaining computational feasibility, we carefully sub-sampled the global sequence dataset. For each subtype, we designed three subsampling strategies to select ∼6000 HA sequences collected from January 2007 to December 2023, of which half (∼3000) were allocated to Southeastern Asia. In the first sub-sampling scheme (even sub-sampling, main analysis), we sub-sampled equal numbers of sequences per sub-location per year (where available) in Southeastern Asia. In temperate regions, we first allocated ∼600 sequences for each sub-region where available, and then sub-sampled equal numbers of sequences per sub-location per year. The second sub-sampling scheme selected sequences proportional to human population. Specifically, we set the number of sequences for each sub-location proportional to the sub-location-specific population size (with a minimum number of 100 sequences per sub-location) in Southeastern Asia; in temperate regions, we first set that number proportional to population size for each sub-region (with a minimum number of 300 sequences per sub-region), and then sub-sample equal numbers of sequences per sub-location per year within each sub-region. We sub-sampled equal numbers per year, because more recent years are overrepresented in sequence numbers compared to previous years (**Fig. 1f, 1j**). Sequences selected in the third subsampling scheme were proportional to the product of population size and influenza positivity rate binned by ISO year. Throughout, among the smallest sub-sampling units (per sub-location per year), more sub-sampling weights were given to sequences with more complete collection dates and higher quality sequences.

**Fig 1.**
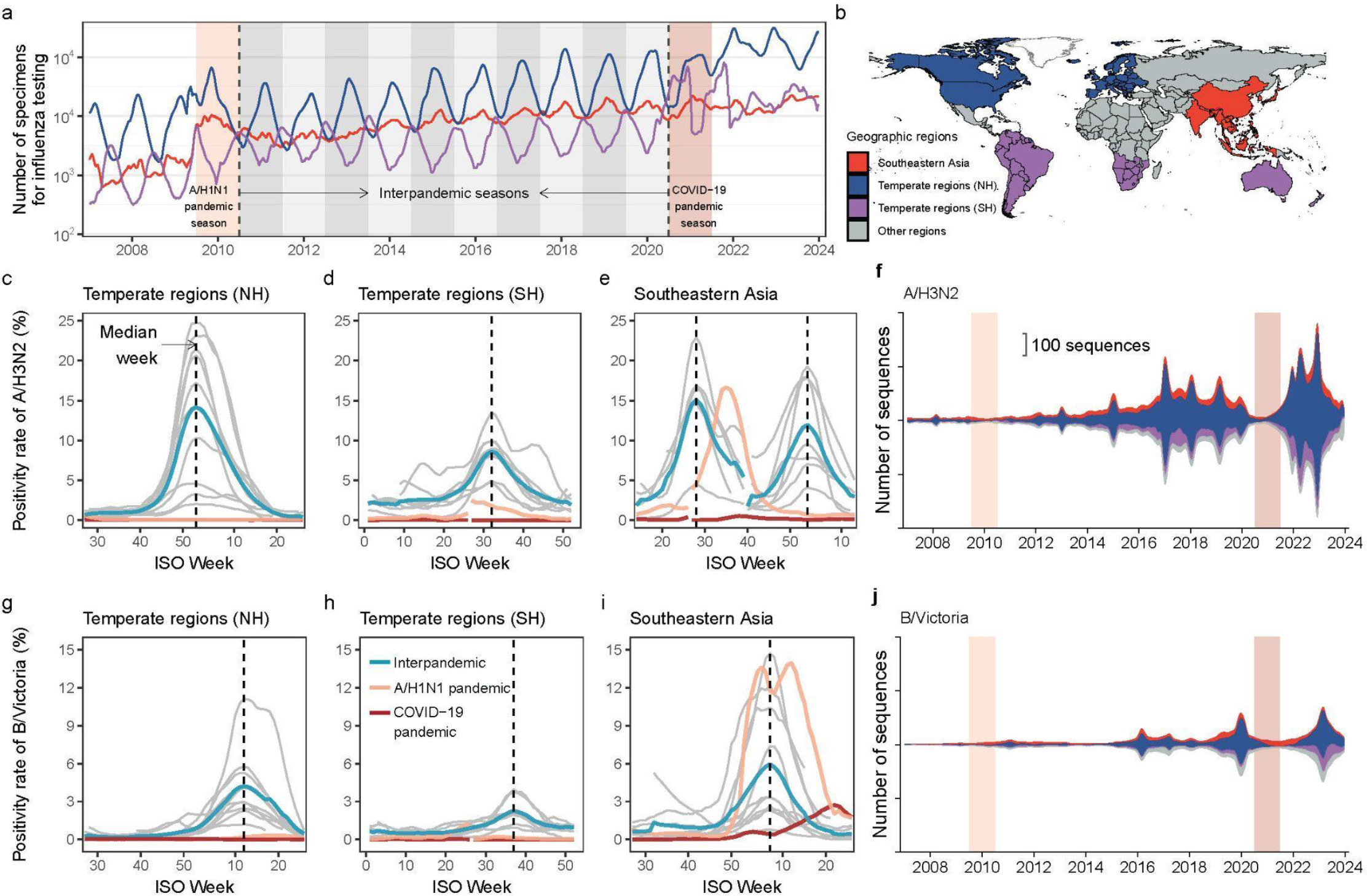
Variation in testing intensity, positivity rate, and numbers of HA gene sequences of seasonal influenza viruses in Southeastern Asia and temperate regions of the northern hemisphere (NH) and southern hemisphere (SH). **a)** A five-week running average of the number of specimens tested for influenza in three geographic regions. Light and dark red shading denote the A/H1N1 and COVID-19 pandemic seasons, respectively, between which are interpandemic seasons. Global-scale data after 2017, presented in **Fig. 1a** of our previous study^10^, have been re-aggregated into three regions and are retained here for the comparisons between the two pandemic seasons. **b)** Geographic divisions used in this study. The entirety of South America was grouped with the southern hemisphere given recommended use of the Southern Hemisphere vaccine formulation throughout the region^5,28^. **c-e)** Average positivity rates for A/H3N2 for seasons in the interpandemic period (cyan-blue lines) versus that during the 2009 A/H1N1 pandemic season (orange lines) and the COVID-19 pandemic season (red lines). Cyan-blue lines represent mean positivity rates for A/H3N2 after aligning the median week of peak (shown as the dashed line) for the seasons in the interpandemic periods, where grey lines show the five-week rolling positivity rate for each season separately after epidemic alignment. The processing details have been presented in **Supplemental Figs. 1-2**. ISO week 53 was removed for some years to maintain temporal consistency. Two epidemic alignments of A/H3N2 were performed in Southeastern Asia by splitting each influenza season into a summer (corresponding to the summer peak) and winter season (corresponding to the winter peak). In panels **d** and **h**, the first half of the positivity lines (Jan-Jun 2010; Jan-Jun 2021) during the pandemic periods occurred after the second half of the line (Jul-Dec 2009; Jul-Dec 2020), for temporal comparison. In panel **e**, the first half of the positivity lines (Apr-Jun 2010; Apr-Jun 2021) during the pandemic periods occurred after the second half of the lines (Jul 2009-Mar 2010; Jul 2020-Mar 2021). **f)** HA gene sequences of A/H3N2 stratified by geographic regions over time. **g-i)** Same as **c-e** but for B/Victoria. **j)** Same as **f** but for B/Victoria.

### Phylogenetic analyses

We aligned sequences in the sub-sampled datasets in Nextclade and only kept the coding regions^20^. We then constructed maximum likelihood (ML) phylogenies using IQ-TREE2^29^. The resulting phylogenetic trees were inspected in TempEst to identify and remove temporal outliers^30^. To better manage the number of sequences, we further reduced clades of sequences where all sequences originated from a single sub-location to a single representative sequence, since such clades contain no information about transitions across sub-locations. This subsampling process resulted in reducing the datasets to ∼4000 genetic sequences.

The ML tree was re-estimated for the reduced genetic datasets using the same specifications described above. We then inferred the time-calibrated tree using TreeTime^31^, which served as the starting tree for Bayesian phylogenetic inference. Phylogenetic trees were inferred in a Bayesian framework using BEAST v1.10.5^32^ and the high-performance BEAGLE library^33^, in which we incorporated a starting ML tree, a HKY nucleotide substitution model with gamma-distributed rate variation among sites, a constant coalescent prior using a

Hamiltonian Monte Carlo (HMC) kernel on the population size and node heights, as well as a strict molecular clock model. Samples with incomplete dates had dates of sampling estimated assuming a uniform prior within the known temporal bounds. These analyses were run for 400-600 million MCMC steps for three chains and sampled every 100,000 steps, with the first 10%-15% considered as burn-in.

### Phylogeographic analyses

#### 1. Two-state phylogeographic analysis

Using the posterior set of trees from the above phylogenetic analyses as a set of empirical trees, we performed a two-state time-inhomogeneous asymmetric discrete trait analysis (DTA) in BEAST 1.10.5, where sequences collected from Southeastern Asia and temperate regions were labelled based on their respective geographic deme. We used the epoch extension to specify five epochs where each epoch spans a single or multiple influenza seasons: i) before the A/H1N1 pandemic season (before 30 June 2009), ii) A/H1N1 pandemic season (from 1 July 2009 to 30 June 2010), iii) interpandemic period (from 1 July 2010 to 30 June 2020), iv) COVID-19 pandemic season (from 1 July 2020 to 30 June 2021), and v) after the COVID-19 pandemic season (after 1 July 2021), enabling us to account for variations in transition rate. Individual counts of transitions between demes were estimated by stochastic mapping in the form of Markov jumps and rewards^34^.

#### 2. Identifying persistent lineages

We identified those Southeastern Asia virus strains that directly descended from the Southeastern-Asia trunk node of the trees, where the trunk was defined as all branches ancestral to viruses sampled within 1 year of the most recent sample^23^. We referred to those lineages as “persistent lineages”, which can be considered as roughly equivalent to opposite of the “transmission lineages” defined in^35^. To achieve this, in brief, we first initiated a depth-first search from each Southeastern-Asia tip for each tree in posterior samples. We defined the Southeastern-Asia tip as belonging to a persistent lineage in this tree if the trunk node from which it descends was associated with “Southeastern-Asia” and also all ancestor nodes in the path from tip to this trunk node were associated with “Southeastern-Asia” as well (**Fig. 3a**). We then summarised the distribution of “persistent lineage” for the posterior set of trees, for which those tips that are classified as belonging to persistent lineages in more than 50% of the posterior tree samples would be ultimately labelled as belonging to “persistent lineages”.

#### 3. Internal GLM-diffusion phylogeographic analysis

The internal movement network within Southeastern Asia was inferred using only sequences classified as belonging to persistent lineages in the prior analysis. We inferred the phylogenetic trees of persistent lineages in BEAST v1.10.5 with a parameter-rich setting where we specified an SRD06 nucleotide substitution model^36^, a Bayesian SkyGrid coalescent prior (with grid points equidistantly spaced in six month intervals)^37^, and a strict molecular clock model. Using the posterior trees from this analysis as empirical trees, we performed a time-inhomogeneous phylogeographic (DTA) model with a generalised linear model (GLM) between the 22 Southeastern Asia sub-locations defined above, with Markov jumps and rewards logged to estimate the transition events^34^. Both overall and relative transition rates were set to be epoch-specific, using the same epochs as above. We collated, aggregated, and standardised time-inhomogeneous airline traffic volumes in the five epochs^10^. This was then used as the sole covariate in the phylogeographic GLM model, assuming time-homogeneous effect sizes and inclusion probabilities^38^. Airline passenger booking data (referred to as airline passenger volumes) from January 2011 to December 2023 were accessed from Official Airline Guide (OAG) Ltd. through a data sharing agreement. Considering that no airline passenger data were available before 2011, we adopted the airline capacity data (the number of seats) from OAG to assume a proportional relationship with the number of passengers travelling^2^ (**Extended Data Figs. 2-3**). Therefore, the airline data of the five epochs refers to i) airline capacity from January 2007 to June 2009; ii) airline capacity from July 2009 to June 2010; iii) airline passenger volumes from January 2011 to June 2020; iv) airline passenger volumes from July 2020 to June 2021, and v) airline passenger volumes from July 2021 to December 2023, respectively.

#### 4. Summary of posterior trees

We used the TreeMarkovJumpHistoryAnalyzer tool to obtain posterior summaries of all Markov jump events from posterior trees^38^. In terms of viral movements from Southeastern Asia to temperate regions, we averaged the weekly movement intensity per season after aligning the movement time series based on the median peak weeks for the seasons during the interpandemic period, with the outlier seasons (e.g., no peak in that season) removed^16,17^. The procedure followed is similar to the methodology illustrated in **Supplemental Fig. 1**.

For A/H3N2 persistent lineages circulating within Southeastern Asia, we estimated the PhyloSor similarity between each pair of three sub-regions (East Asia, South Asia, and Southeast Asia) in Southeastern Asia, which quantifies the similarity of viral populations between those locations as the proportion of branch lengths in phylogenetic trees that are shared relative to the total branch lengths of both populations^39^. Additionally, we computed Euclidean distances among seasons for those vectorized asymmetric jump matrices among three sub-regions (East Asia, South Asia, and Southeast Asia), and then performed a classical multidimensional scaling in a 2-dimensional space^40^. Finally, we summarised the trunk location within Southeastern Asia over time based on the phylogeographic estimates using PACT v.0.9.5 (https://github.com/trvrb/PACT)^23^.

### Sequence-based antigenic distance

As the antigenic map has been clearly resolved for A/H3N2^41^, we calculated sequence-based antigenic distances for A/H3N2 subtypes following a genotype-phenotype mapping^42^. In brief, in each antigenic site (A, B, C, D, E) of A/H3N2^41^, we quantify the number of differences in amino acids (Hamming distance) for aligned amino acid sequences compared to the sequence of the vaccine strain “A/Wisconsin/67/2005” (also the reference sequence in this study). Antigenic site-specific Hamming distances are divided by the total number of amino acids in the antigenic site, and then multiplied by 20, representing a 20-dimensional immunological shape space^43^. The final antigenic distance between each virus strain and “A/Wisconsin/67/2005” was calculated by averaging the above values at five antigenic sites. To identify the leading and trailing geographic regions undergoing antigenic evolution of A/H3N2, we fitted a linear best-fit line between antigenic distance against date of collection. Points to the right of the line are thought to be antigenically advanced, whereas strains to the left of the line are antigenically lagging^4^. We then summarised the leading and trailing pattern by regions and time periods. Bootstrap resampling of distance values yielded a *p* value for the difference between interpandemic seasons and each pandemic season. We could not perform the same analysis for influenza B/Victoria due to the less resolved mapping between genotype and antigenic phenotype.

## Results

### Disruptions of influenza activity during pandemic seasons

To synchronise with the northern hemisphere influenza seasons (from July 1st of one year to June 30th of the next year^22,44^), the A/H1N1 and COVID-19 pandemic periods were respectively defined as spanning from July 2009 to June 2010 and from July 2020 to June 2021, with the periods between them labelled as the interpandemic periods/seasons. Similar to our previous work relating specifically to the COVID-19 pandemic period^10^, the sampling intensity of virological surveillance for seasonal influenza during the two pandemic seasons was no lower than during previous seasons (**Fig. 1a**). The amplitude of surveillance intensity fluctuated across region and time, with a large increase directly after the start of the A/H1N1 pandemic and a generally increasing trend thereafter **(Fig. 1a)**. Yearly fluctuations in viral sampling were observed in temperate regions, peaking in their respective winter months **(Fig. 1a-b)**. On the other hand, surveillance intensity within any given year remained relatively stable in Southeastern Asia (**Fig. 1a-b**), due to persistent influenza circulation throughout the year in large parts of the region^45,46^.

Given the emergence of A/H1N1pdm09 in 2009^18^ and the subsequent replacement of the previously circulating seasonal A/H1N1 virus, as well as the potential disappearance of B/Yamagata in 2020^19^, we focused on the remaining two human influenza subtypes, A/H3N2 and B/Victoria, for which we could study disruptions associated with two pandemic seasons. Initially, we established the average curve of positivity rate (as a proxy of influenza activity^17^) for seasons in the interpandemic period following epidemic alignment by peak week (details in **Methods**) as a baseline, against which the positivity rates during the two pandemic seasons were contextualised (**Fig. 1c-e, 1g-i**).

In temperate regions we observe a single annual winter wave of H3N2 circulation during the interpandemic period, while biannual peaks are observed in Southeastern Asia. Compared to interpandemic averages, extremely low positivity rates were observed in all three regions throughout the two pandemics, except for a single peak occurring in Southeastern Asia in late August 2009 during the A/H1N1 pandemic, without a subsequent second peak (**Fig. 1c-e**). This single A/H3N2 wave was hypothesised to be associated with limited implementation of NPIs during the A/H1N1 pandemic^47^, while the absence of a second peak could be attributable to viral interference due to large scale transmission of the novel A/H1N1 pandemic virus^48^, presumably via heterosubtypic cross-immunity^6^. The low positivity rate of A/H3N2 during the COVID-19 pandemic in all regions further highlights the impact of human behavioural changes on influenza circulation.

Despite variations in B/Victoria activity from season to season, we observe a single peak in all three regions during the interpandemic period, with a lower and usually delayed peak compared to A/H3N2 (**Fig. 1g-i**). A larger than usual wave of B/Victoria occurred in Southeastern Asia in the first quarter of 2010 during the A/H1N1 pandemic, whereas little circulation was observed during the COVID-19 pandemic (**Fig. 1i**). We hypothesise that this difference can be partly explained by varying intensity and duration of NPIs implemented during the two pandemics, with a 70.9% decrease of global airline traffic in 2020 compared to 2019, compared to a 2.4% reduction from 2008 to 2009 (**Extended Data Fig. 2a**).

### Impact of pandemics on influenza movements from Southeastern Asia to temperate regions

Given the heterogeneous spatiotemporal distribution of genetic sequences (**Fig. 1f, 1j**), we adopted three sub-sampling schemes to select sequences and assess the potential impact of sampling biases. All three sub-sampling schemes resulted in similar numbers of Markov jump counts between locations over time, for both subtypes (**Extended Data Fig. 4**), indicating that the signal is robust to the sub-sampling scheme. Thus, in our main analysis we arbitrarily employed an even sub-sampling scheme by time and location (details in **Methods**). We first extended a previous study^10^ by performing a two-state discrete trait analysis to estimate Markov jump events (referred to as viral movement events) between Southeastern Asia and temperate regions from 2007 to 2023 (**Extended Data Fig. 4, Fig. 2**). Despite selecting a similar number of genetic sequences from both regions, a higher frequency of A/H3N2 movements from Southeastern Asia to temperate regions was detected, compared to the reverse direction (**Extended Data Fig. 4a, Fig. 2a**). Regarding B/Victoria lineages, the pattern of bidirectional movements between the two regions varied widely from season to season (**Extended Data Fig. 4d**), but with a more balanced flux than A/H3N2 (**Fig. 2a-b**).

**Fig 2.**
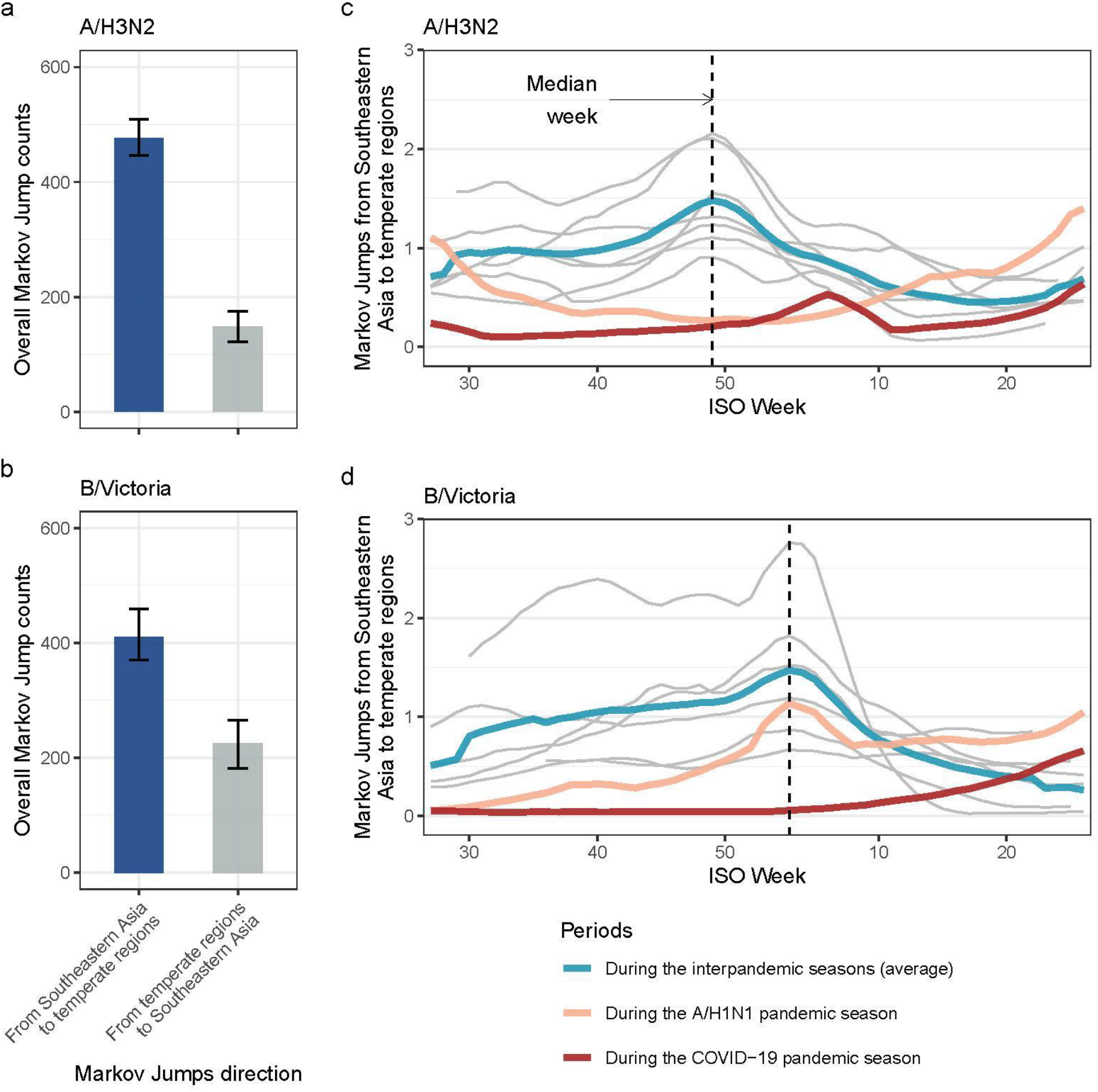
Inferred movement of A/H3N2 and B/Victoria between Southeastern Asia and temperate regions. **a-b)** The overall Markov Jump counts in two directions over the entire time period of interest (A/H1N1 pandemic season, interpandemic seasons, and COVID-19 pandemic season). The error bar indicates the 95% HPD interval. **c-d)** Average Markov Jumps of A/H3N2 and B/Victoria from Southeastern Asia to temperate regions for the seasons in the interpandemic period (after removing outlier seasons and aligning at the median peak). Grey lines show Markov Jumps for each season in the interpandemic period after epidemic alignment; cyan is the interpandemic average, orange the A/H1N1 pandemic season and red the COVID19 pandemic season. ISO week 53 was removed for some years to maintain temporal consistency.

Subsequently, we averaged the Markov jump counts from Southeastern Asia to temperate regions for the seasons during the interpandemic period, after epidemic alignment at median week for timing of each season. As a baseline, A/H3N2 movement from Southeastern Asia gradually peaked in early December in interpandemic seasons (**Fig. 2c**), while B/Victoria movements peaked in January, although with a heavy tail toward earlier weeks, indicating sustained movements starting much earlier **(Fig. 2d)**. During the two pandemics, the typical A/H3N2 winter peak in exports from Southeastern Asia disappeared. During the A/H1N1 pandemic, B/Victoria movement peaked at the same time as during the inter-pandemic period, and at a level consistent with inter-pandemic activity. This coincides with the large B/Victoria wave in Southeastern Asia during the same time period, likely resulting in sustained exports from Southeastern Asia, but not establishment in temperate regions (**Fig. 1g-i**). However, no corresponding B/Victoria winter peak was detected during the COVID-19 pandemic.

### Persistent influenza lineages within Southeastern Asia

To enable further reconstruction of the internal network within Southeastern Asia, we categorised the viruses circulating in Southeastern Asia based on whether they had been inferred to have either been newly introduced or persisted in the region from a previous season. Briefly, for lineages in Southeastern Asia, “persistent lineages” are defined as those lineages descended from the most recent Southeastern Asia trunk node. Although we do not interpret this in terms of epidemiological persistence of transmission chains between seasons, these lineages can be seen as more likely to have persisted from previous seasons rather than being recently introduced into Southeastern Asia (**Fig. 3a**, details in **Methods**).

**Fig 3.**
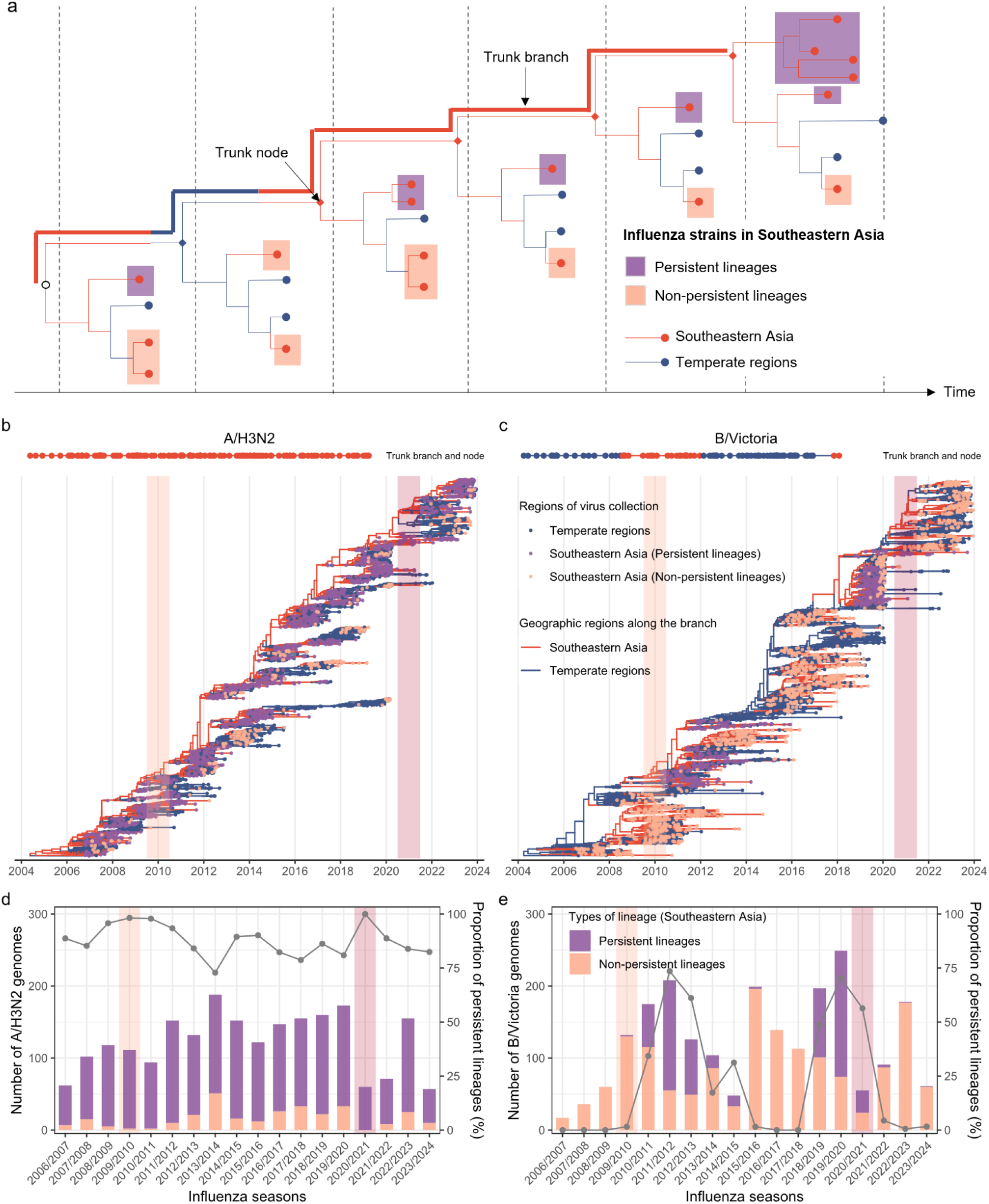
Persistent lineages within Southeastern Asia and their topological and temporal distributions. **a)** Schematic diagram illustrating how viruses circulating in Southeastern Asia belonging to persistent lineages were identified. A Southeastern-Asia tip was defined as belonging to a persistent lineage if its nearest trunk node was associated with “Southeastern-Asia” and all ancestor nodes from this tip to the nearest trunk node were associated with “Southeastern-Asia” backwards in time. Trunk was defined as all branches ancestral to viruses sampled within 1 year of the most recent sample. **b-c)** The maximum clade credibility (MCC) tree of A/H3N2 and B/Victoria lineages. Tips are coloured by geographic region of virus collection, in which strains in Southeastern Asia are separately annotated as belonging to persistent and non-persistent lineages; internal branches are coloured by geographic region as inferred by Bayesian phylogeographic methods. Trunk nodes over time are presented as dots coloured by trunk location on a horizontal line at the top of the panels. Trunk assignments are summarised up to the most recent common ancestor (tMRCA) of viruses sampled within 1 year of the most recent sample. **d-e)** The temporal distribution of the numbers of sub-sampled virus genomes from Southeastern Asia belonging to persistent and non-persistent lineages over influenza seasons, for A/H3N2 and B/Victoria lineages, respectively (left y-axis). The proportion of virus genomes belonging to persistent lineages over time is represented by the grey line (right y-axis).

The maximum clade credibility (MCC) tree of A/H3N2 exhibited a trunk that was relatively consistently associated with Southeastern Asia, whereas the trunk location of B/Victoria displayed variations across influenza seasons (**Fig. 3b-c**), consistent with more balanced B/Victoria fluxes between Southeastern Asia and temperate regions (**Fig. 2a-b**). This aligns with past observations that global A/H3N2 circulation patterns exhibit a stronger and more predictable source-sink dynamic than B/Victoria^4,23,49^. We found that a higher percentage of A/H3N2 viruses sequenced in Southeastern Asia belonged to persistent lineages (86.5%, 1913/2211) than B/Victoria viruses (29.1%, 636/2188) (*P* < 0.001, chi-square test), consistent for both the pandemic and interpandemic periods (**Fig. 3d-e**). Interestingly, the percentage of A/H3N2 viruses in Southeastern Asia belonging to persistent lineages appears to have increased during both pandemic seasons (**Fig. 3d**), possibly due to the reduction of human mobility and subsequent lineage movement.

### Shifts in antigenically leading and trailing patterns

To provide more insights into the evolutionary heterochrony across regions and seasons, we calculated sequence-based antigenic distance for A/H3N2, a subtype whose antigenic map has been clearly resolved^41^. Based on the HA genetic sequence of A/H3N2, we calculated the Hamming distances of each virus strain to the vaccine strain (A/Wisconsin/67/2005) across the five major antigenic sites following the approach from^42^. We found that A/H3N2 viruses evolved linearly from 2007 to 2023 (**Fig. 4a**), with no clear outliers during the pandemic seasons. This suggests that A/H3N2 viruses circulating in both Southeastern Asia and temperate regions underwent antigenic evolution in a globally homogeneous way, consistent with the ladder-like tree topology of A/H3N2 (**Fig. 3a**) and previous findings^4,50^.

**Fig 4.**
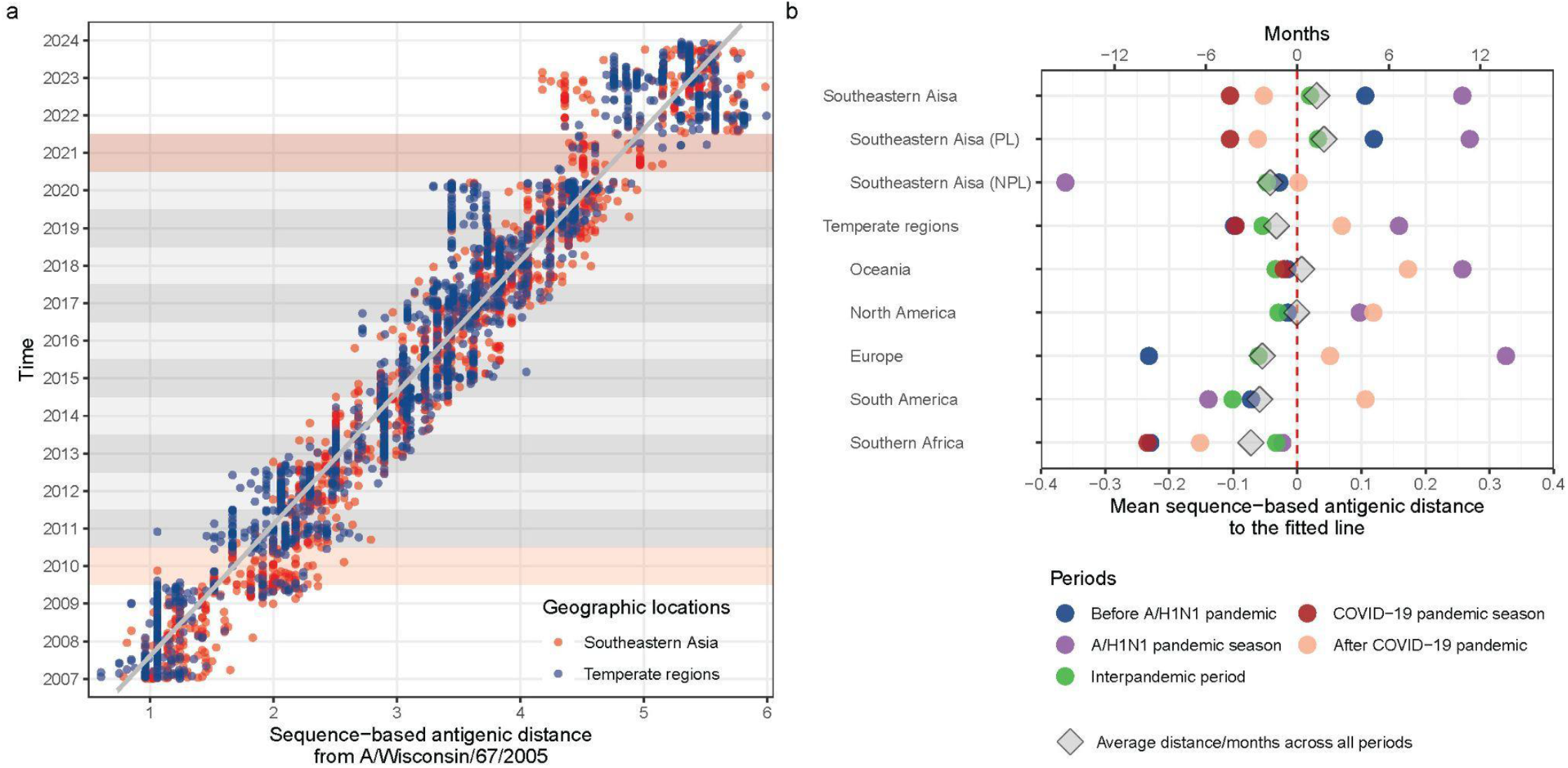
Leading and trailing geographic regions of A/H3N2 antigenic evolution. **a)** Sequence-based antigenic distance from A/Wisconsin/67/2005 of all A/H3N2 virus strains plotted against time of collection. The thick grey line is the fitted line using a linear model. Points to the right of the line are antigenically advanced, whereas strains to the left of the line are antigenically lagging. Light and dark red shading denotes the A/H1N1 and COVID-19 pandemic seasons, respectively. **b)** Antigenic distance to the fitted line by region and period. Grey oblique squares indicate the average antigenic distance to the fitted line for strains isolated in each region. Coloured circles split this overall average by various periods. Points to the right of the graph are antigenically advanced, whereas strains to the left are antigenically lagging. Antigenic distance can also be interpreted as time using the slope of the regression line in panel **a**; thus, time is shown as a second x axis (top). The absence of observations in certain periods is attributed to incomplete gene sequences, making it challenging to compute antigenic distance. PL, persistent lineages; NPL, non-persistent lineages.

Next, we explored which regions typically lead or lag A/H3N2 antigenic evolution, and whether pandemic events may have disrupted these patterns. We computed the distance of each strain to the fitted line between antigenic distance and time of collection, for which points to the right of the line indicated antigenically advanced strains, while those to the left indicated antigenically lagging strains^4^ (**Fig. 4a**). During the interpandemic period, newly emerged A/H3N2 strains appeared in Southeastern Asia on average, 3.1 months earlier than in temperate regions, while the greatest lag in arrival of antigenic novelty was found in South America with a delay of 5.1 months (**Fig. 4b**). Within Southeastern Asia, persistent A/H3N2 strains appeared on average 3.3 months earlier than non-persistent lineages. Additionally, this leading and trailing pattern differed from period to period (**Fig. 4b**). During the A/H1N1 pandemic, newly emerged A/H3N2 strains appeared on average 10.0 months earlier in Southeastern Asia and 9.0 months earlier in temperate regions compared to the interpandemic period (both *P* < 0.001, **Extended Data Table 1**). However, during the COVID-19 pandemic, A/H3N2 antigenic evolution lagged in both Southeastern Asia (by 5.3 months, *P* < 0.001) and temperate regions (by 1.8 months, *P* = 0.632) compared to the interpandemic period (**Extended Data Table 1**). The time-advanced pattern of A/H3N2 antigenic evolution seen globally during the A/H1N1 pandemic might be linked to modest NPIs and enhanced natural selection from competition with high circulation of A/H1N1pdm09 worldwide. Conversely, the lagged pattern of A/H3N2 evolution seen in Southeastern Asia during the COVID-19 pandemic could be attributed to limited influenza circulation and evolution due to human behaviour changes driven by stringent and long-lasting NPIs. The non-significant differences of antigenic evolution during the COVID-19 pandemic season in temperate regions (*P* = 0.632) might be associated with earlier re-opening of these regions (**Extended Data Fig. 5**), and the fact that these regions do not typically lead antigenic novelty. More population-based and modelling studies are required to deepen the understanding of the exact mechanisms underpinning our observations.

### Characterising the A/H3N2 movement network within Southeastern Asia

Next, we used the persistent lineages of A/H3N2 within Southeastern Asia, identified earlier, to reconstruct the transmission network in this region. Although the inferred internal network fluctuates from season to season, we constructed an average network using the intensity of viral movement between pairs of sub-locations per season during the interpandemic period as a baseline, and contrasted baseline patterns with those of pandemic seasons (**Fig. 5a**). During the A/H1N1 pandemic, the overall number of inferred virus movement events declined by 52.6% (1-85.0/179.4) compared to the baseline, whereas movement intensity increased for routes from Indonesia-East Timor (8.5 vs 5.7 in the baseline) and Laos (3.7 vs 2.6) (**Fig. 5b, Extended Data Fig. 6**). During the COVID-19 pandemic, the overall number of viral movements declined by 86.1% (1-25.0/179.4) compared to the interpandemic period (**Fig. 5c**). The reductions were consistent for the epoch-specific dispersal rates accounting for the tree length (decreased by 54.7% and 79.2% during the 2009 A/H1N1 and COVID-19 pandemic seasons, respectively) (**Fig. 5d**). We further subdivided Southeastern Asia into East Asia, South Asia, and Southeast Asia to perform a multidimensional scaling analysis for vectorized asymmetric movement matrices among these three sub-regions. There was a marked divergence in internal movement network structure during the COVID-19 pandemic, and to a lesser extent during the A/H1N1 pandemic season, in comparison to the well-mixed pattern that was observed during the interpandemic seasons (**Fig. 5e**). Further analysis of the trunk location revealed a temporally varying pattern (**Extended Data Fig. 7**), indicating that the internal network within Southeastern Asia is a dynamic process that is maintained not only by one sub-location as the source population, consistent with prior work^4^.

**Fig 5.**
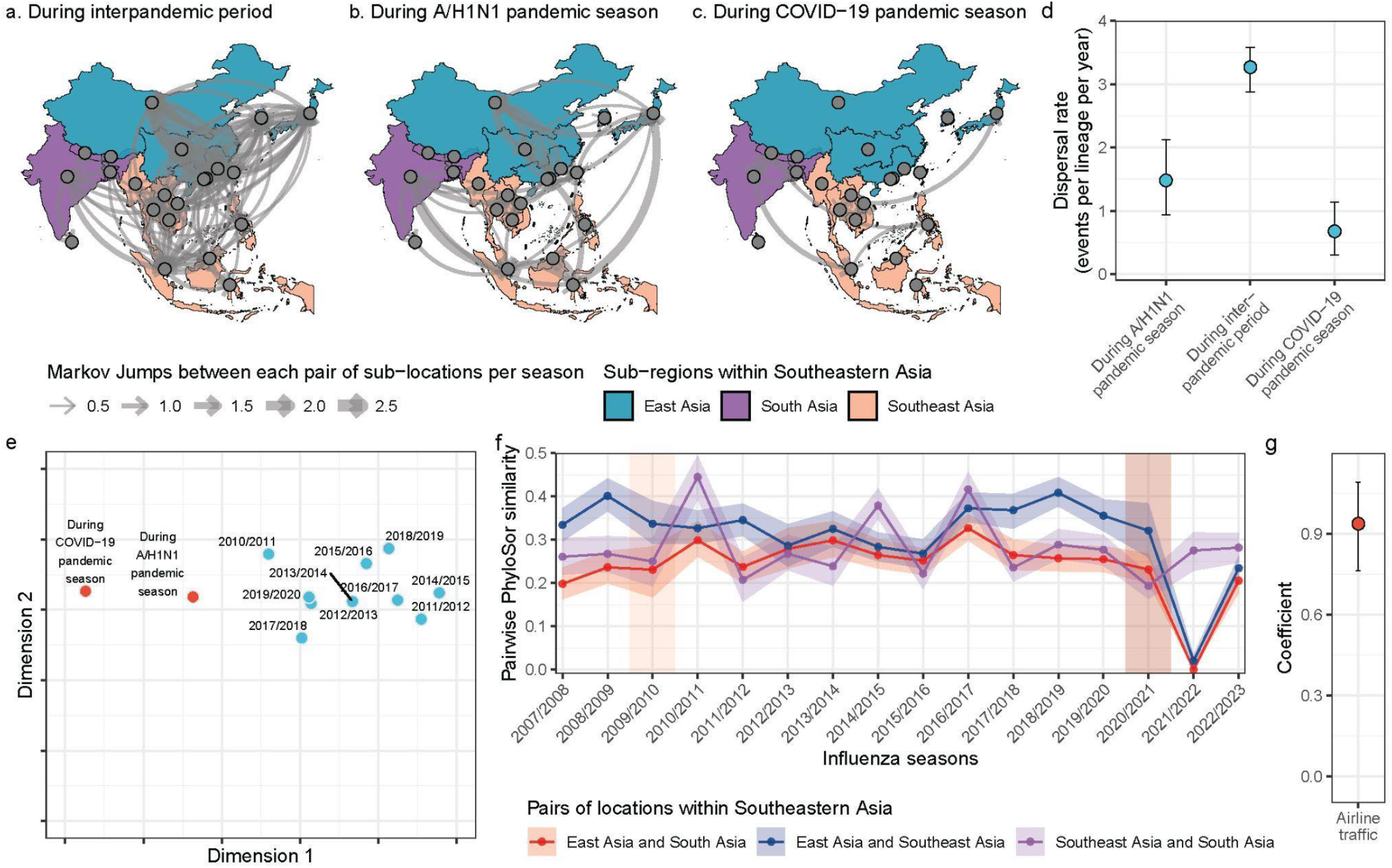
The migration network, phylogenetic similarity, and dispersal drivers of persistent A/H3N2 strains within Southeastern Asia. **a-c)** The average Markov Jump counts between pairs of sub-locations per season during the three periods. Only lines representing a number of Markov Jumps greater than 0.5 per season are shown, for clarity. Three sub-regions within Southeastern Asia were coloured, where East Asia only encompasses China, Japan, and South Korea, while South Asia only includes Bangladesh, Bhutan, India, Sri Lanka, and Nepal. **d)** Period-specific dispersal rates inferred during the phylogeographic reconstruction. **e)** Multidimensional scaling analysis for vectorized asymmetric jump matrices among three sub-regions (East Asia, South Asia, and Southeast Asia) in different influenza seasons. **f)** Pairwise PhyloSor similarity of virus strains circulating between pairs of sub-regions within Southeastern Asia over influenza seasons as a proxy for lineage mixing. The smaller the Pairwise PhyloSor similarity is, the less mixed the lineages circulating between pairs of sub-regions are. The shaded area around the line indicates 95% HPD interval. **g)** Contributions of airline traffic to internal diffusion of A/H3N2 in Southeastern Asia. The error bar indicates the 95% HPD interval.

We further estimate the A/H3N2 phylogenetic similarity using the tree topology and branch length^39^ (PhyloSor, details in **Methods**), as a proxy for lineage mixing among the three sub-regions (East Asia, South Asia, and Southeast Asia). During the A/H1N1 pandemic season, there was a considerable amount of lineage mixing between pairs of sub-regions within Southeastern Asia, aligned with that of prior seasons, but a notable increase was seen in 2010-2011 between Southeast Asia and South Asia (similarity: 0.44, 95% highest posterior density (HPD): 0.39 to 0.49, **Fig. 5f**). The extent of lineage mixing only slightly declined during the COVID-19 pandemic season in 2020-2021 for all pairs of sub-regions, but declined precipitously in 2021-2022 between East Asia and South Asia, and East Asia and Southeast Asia. We hypothesise that virus lineages are still able to circulate on the same branch of a tree phylogeny despite low activity, explaining the “delayed” effect of the pandemic on A/H3N2 similarity. The fact that during the 2021/2022 season persistent lineages in East Asia rarely mixed with those from the other two sub-regions, while mixing was at typical levels between Southeast Asia and South Asia (**Fig. 5f**) could be attributed to the uniquely stringent NPIs and international travel restrictions that were still in place in China over this period. Thereby lineages from East Asia had limited opportunities to mix with lineages from other regions (**Extended Data Fig. 3**). This observation is further supported by the significant contribution of airline traffic (coefficient: 0.94, 95% HPD: 0.76 to 1.09) to A/H3N2 spread within the whole Southeastern Asia region (**Fig. 5g**).

## Discussion

We expanded and refined our previous work^10^ by focusing on Southeastern Asia and comparing two distinct pandemic events at a finer spatiotemporal resolution. We demonstrated that typical Autumn-Winter waves of influenza lineage movements from Southeastern Asia to temperate regions disappeared almost completely during both pandemic-related disruptions, with the exception of B/Victoria lineages during the A/H1N1 pandemic. However, while we observed a wave of B/Victoria exports from Southeastern Asia to temperate regions during the A/H1N1 pandemic, coinciding with a large B/Victoria epidemic wave in Southeastern Asia, these exports did not spark B/Victoria epidemic waves in temperate regions, suggesting little to no establishment of exported lineages. We found opposite patterns of pandemic-related disruptions in A/H3N2 antigenic evolution, with a greater lead time during the 2009 A/H1N1 pandemic, and a greater lag time during the COVID-19 pandemic, compared to inter-pandemic patterns. Additionally, a higher proportion of A/H3N2 lineages circulating within Southeastern Asia have been estimated to persist across seasons as compared to B/Victoria lineages. By analysing those persistent lineages, we highlight how the A/H3N2 internal migration network within Southeastern Asia is characterised by a dynamic change in trunk locations over time. During the COVID-19 pandemic season, it was shaped by reductions in the intensity of inferred viral movements, the diverse structure of the internal movement network, and decreased lineage mixing. Changes in the circulation network within Southeastern Asia were less pronounced during the A/H1N1 pandemic season likely due to fewer pandemic related disruptions at that time.

We extended previous work^4,23,49^ to consider a broader geographic scope of Southeastern Asia and characterise the main trunk location for the global influenza circulation. Our data suggest that Southeastern Asia plays a more conspicuous role in the global circulation of A/H3N2 compared to B/Victoria. The importance of Southeastern Asia in the evolution of A/H3N2 has been further attributed to the seasonal nature of influenza in temperate regions, in which strong genetic and transmission bottlenecks lower the likelihood of local persistence and global fixation of circulating strains^14,51^. However, the internal network maintaining the leading role of Southeastern Asia has rarely been explored in a quantitative manner^4^. Our quantitative analyses reveal that the movement network within Southeastern Asia is characterised by highly-connected dispersal routes among the major airline transportation hubs (**Extended Data Fig. 6a**) and a temporally varying trunk location (**Extended Data Fig. 7**). In line with a previous hypothesis^4^, this network of asynchronous but temporally overlapping epidemics, connected by proximity and human mobility, promotes antigenic evolution and global dispersal of A/H3N2 strains. In contrast, global B/Victoria circulation exhibits less dependence on Southeastern Asia, with a pattern of geographically separate evolution and circulation rather than global dispersion, consistent with a previous study^23^. Although the underlying mechanisms driving the heterogeneity across viruses remain unclear, we speculate that the more rapid antigenic drift for A/H3N2 compared to B/Victoria^52^ could potentially result in a greater intrinsic fitness advantage. Additionally, it has been suggested that younger ages of infection for B/Victoria compared to A/H3N2 could lead to less spatial dissemination as children are less mobile than adults^23,53^.

Influenza circulation patterns are also subject to alteration via changes in human behaviour, especially mobility, during pandemics. The occurrence of two global pandemics (2009 A/H1N1 and COVID-19) in the 21st century has provided a natural experiment for evaluating changes in human mobility and immunity and their impact on influenza transmission at the population level (we did not consider other large epidemics/outbreaks, such as Zika virus, as they were localised to specific parts of the world). Understanding of circulation patterns during these two pandemics allows for enhancing awareness and preparedness against influenza outbreaks in inter-pandemic seasons and when facing the next public health emergencies. Differences in the circulation and evolutionary patterns of influenza viruses during the two pandemics can be interpreted by a complex interaction between local NPIs, regional and global mobility, and viral interference. It is evident that the intensity and duration of NPIs during the COVID-19 pandemic were far greater in comparison to the 2009 A/H1N1 pandemic, as partly illustrated by the extent of reduction in airline flight data relative to the pre-pandemic year (**Extended Data Figs. 2-3**). In addition, human behavioural changes including contact reduction and mask wearing will have also contributed to the decline of respiratory infections during the COVID-19 pandemics^54^. Accordingly, perturbation of influenza virus circulation was more drastic during the COVID-19 pandemic, coinciding with our findings where most internal transmission within Southeastern Asia almost halted. Although we could not study the B/Yamagata lineage in this work, the global disappearance of this lineage from surveillance data since late 2020 attests of the marked perturbation of the COVID-19 pandemic period on influenza dynamics^55^.

The novelty of our approach is to contrast the pronounced perturbation of the COVID-19 pandemic with a pandemic of a more moderate kind, the 2009 A/H1N1 pandemic, which we hypothesised would have also affected circulation patterns of resident influenza strains. The novel virus responsible for the 2009 A/H1N1 pandemic is thought to display viral interference with seasonal influenza A virus, presumably via more specific heterosubtypic cross-immunity^6,56^. Since A/H3N2 viruses are more closely related to A/H1N1pdm09 viruses than B/Victoria viruses, massive circulation of A/H1N1pdm09 in the first pandemic wave may have limited the circulation and diffusion of A/H3N2 viruses via competition for susceptibles, in line with a consistent globally negative correlation between influenza A/H3N2 and A/H1N1pdm09 activity^48^. This competition was further illustrated in our analysis by the disappearance of the second A/H3N2 peak wave and A/H3N2 movement wave from Southeastern Asia to temperate regions during the A/H1N1 pandemic season. In contrast, we see little changes in influenza B circulation, consistent with less or no cross-immunity. NPIs could have also played a role in the A/H1N1 pandemic disruptions we evidenced in our study, although NPIs (e.g., school closures) were limited to a few countries and the early weeks of the pandemic^57^. Lastly, variations in pre-existing population immunity prior to the onset of a pandemic might also contribute to differences in influenza circulation observed during two pandemic periods ^58^. Disentangling the roles of NPIs, viral interference, immunity, and other potential factors in shaping the circulation patterns of seasonal influenza merits more investigation.

### Limitations

Our results should be interpreted in the context of several limitations. First, inherent biases exist due to the nature of genetic data from heterogeneous genomic surveillance efforts and data sharing initiatives worldwide. To address this issue we carefully downsampled genetic data geographically and temporally; our main conclusions are robust across various sets of genetic data from different sub-sampling schemes (**Extended Data Figs. 4 and 8**). Nevertheless, we acknowledge that our dataset completely omits data from large and populous parts of the world (e.g., West, Central and East Africa, Central America, Russia etc.) that could potentially play important roles in the global influenza network. However, our aim is not to reconstruct the global spread of seasonal influenza viruses, but rather to focus on the network within Southeastern Asia and the role of Southeastern Asia in seeding epidemics in temperate regions. Secondly, we recognize that the implementation and lifting of NPIs varied across locations and time, and thereby the impact of these changes on influenza circulation patterns may be heterogeneous – here we concentrate on regional averages. For simplicity, our study considered the same pandemic period definition for all countries although NPIs were put in place in a heterochronous way. In addition, the influenza season defined in this study did not uniformly align well with that in the Southern Hemisphere due to the asynchronous influenza circulation worldwide. Third, epidemic alignment of influenza activity and movement across seasons could be affected by some atypical seasons, but we believe that the average multi-year curves enable us to capture baseline seasonality patterns. Lastly, our study only retrospectively tracked the patterns of influenza circulation over a long-term period, without dissecting the contribution of behavioural and immunological factors. However, we believe that the present findings could guide more causal analyses in the future.

### Future perspectives

Our findings may be instructive in improving the global influenza surveillance, intervention, and vaccination strategies through insights from i) different antigenically advanced and lagged patterns across regions and periods, ii) distinct source-sink dynamics emanating from and within Southeastern Asia by virus types and periods, and iii) differential extent of interactions between pandemic disruption and viral dynamics. Temporally varying networks within Southeastern Asia further support the need for geographically extensive virological and genomic surveillance in this region, allowing for earlier identification of antigenically novel strains and thereby providing a longer lead time for developing and rolling out well-matched vaccines. In addition to the need for forecasts of influenza evolution and epidemic size^59,60^, establishing a global influenza migration network at a finer geographic scale (e.g., country level; over 120 WHO Member States have contributed to influenza surveillance as of May 2024^15^) also holds promise for optimising vaccine recommendations tailored to each country’s future influenza outbreaks.

## Conclusions

In summary, our study provides a comprehensive reconstruction of the circulation patterns of seasonal influenza viruses both out of and within Southeastern Asia during the 2009 A/H1N1 and COVID-19 pandemic with comparison to the interpandemic period. We highlighted the heterogeneous impact of two distinct pandemic-related disruptions on evolution and mixing of seasonal influenza, focusing on the transmission dynamics within Southeastern Asia. Our empirical findings can help anticipate the effects of adopting control measures as routine practices to mitigate the disease burden of seasonal influenza, as well as preparedness against outbreaks of influenza or other seasonal respiratory pathogens in future pandemic scenarios.

## Data availability

Influenza virological surveillance data were available from FluNet (https://www.who.int/tools/flunet). Genetic sequences used were downloaded from NCBI (https://www.ncbi.nlm.nih.gov/labs/virus/vssi/<u>#/</u>) and GISAID (https://www.gisaid.org/). The origin-destination air flight data were provided by Official Airline Guide (OAG) Ltd. (https://www.oag.com/) through a data sharing agreement.

## Data Availability

All data produced in the present study are available upon reasonable request to the authors

## Acknowledgements

We gratefully acknowledge all data contributors, i.e., the Authors and their Originating laboratories responsible for obtaining the specimens, and their Submitting laboratories for generating the genetic sequence and metadata and sharing via the GISAID Initiative and NCBI, on which this research is based. The acknowledgment table of genetic data used is provided on our GitHub repository. The computations in this research were performed using the CFFF platform of Fudan University. H.Y. acknowledges financial support from the Key Program of the National Natural Science Foundation of China (No. 82130093) and the General Program of the National Natural Science Foundation of China (No. 82073613). M.U.G.K. acknowledges funding from The Rockefeller Foundation, Google.org, the Oxford Martin School Pandemic Genomics programme, European Union’s Horizon Europe programme projects MOOD (#874850) and E4Warning (#101086640), the John Fell Fund, a Branco Weiss Fellowship and Wellcome Trust grants 225288/Z/22/Z, 226052/Z/22/Z & 228186/Z/23/Z, United Kingdom Research and Innovation (#APP8583) and the Medical Research Foundation (MRF-RG-ICCH-2022-100069). P.L. acknowledges support from the European Research Council (grant agreement no. 725422 – ReservoirDOCS) and from the Research Foundation - Flanders (‘Fonds voor Wetenschappelijk Onderzoek - Vlaanderen’, G0D5117N, G005323N and G051322N). Z.C. acknowledges financial support from the National Natural Science Foundation of China (No. 823B2089). J.C. acknowledges financial support from the Young Scientists Fund of the National Natural Science Foundation of China (No. 82304199). The contents of this publication do not necessarily reflect the views of the funders.

## Author contributions

H.Y. conceived and planned the research. Z.C. and P.L. analysed the data. J.L.H.T., J.C., C.V., L.D.P., P.L., M.U.G.K. and H.Y. advised on methodologies. Z.C. wrote the initial manuscript draft. Z.C., C.V., L.D.P., P.L., M.U.G.K. and H.Y. interpreted the results and revised the content critically. All authors edited, read, and approved the manuscript.

## Disclaimer

The findings and conclusions in this report are those of the authors and do not necessarily represent the official position of the US National Institutes of Health or Department of Health and Human Services.

## Extended data

**Extended Data Fig 1.**
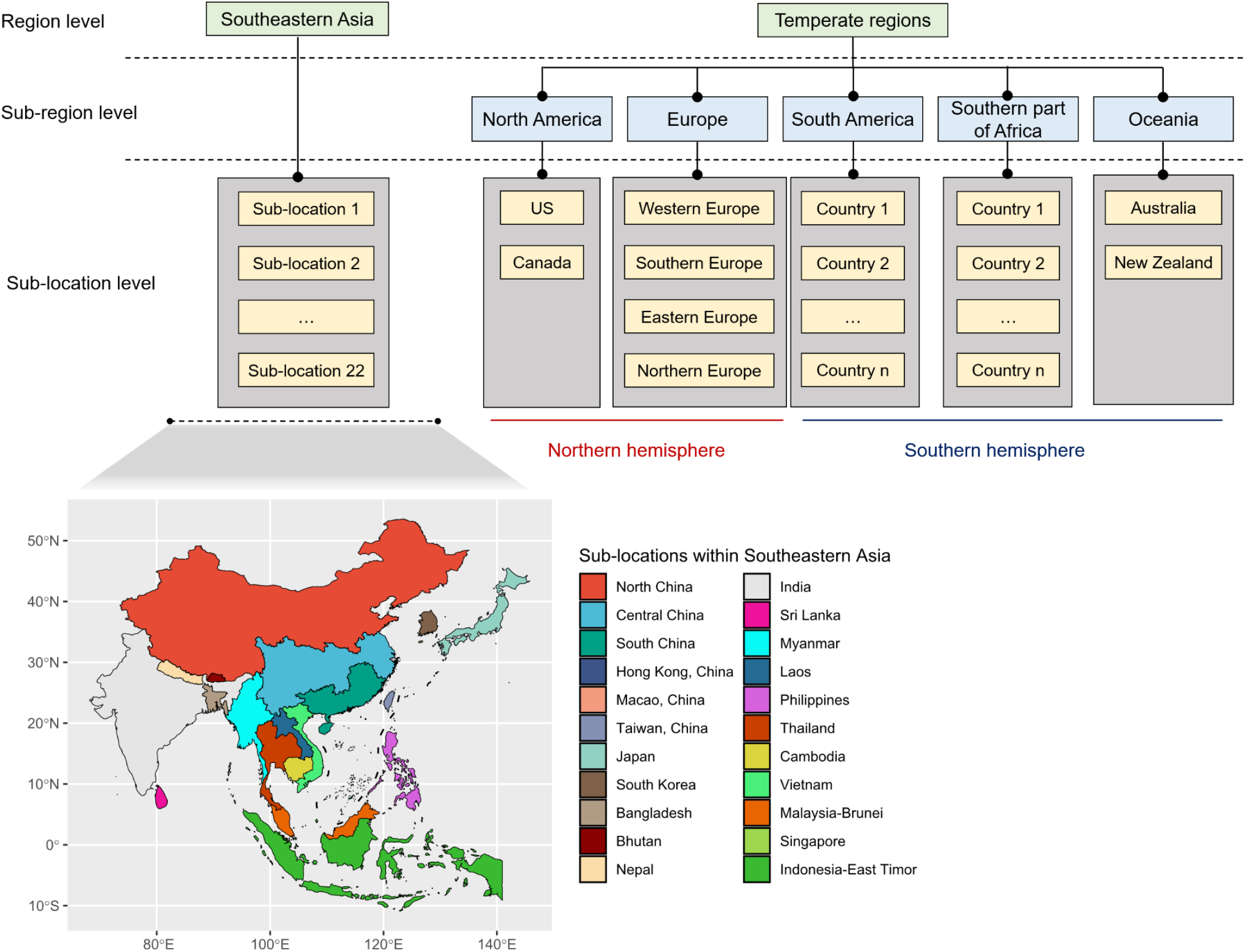
The hierarchical geographic structures used for the sub-sampling process. The smallest sub-sampling unit was in the sub-location level. The entirety of South America was grouped with the southern hemisphere (SH), since the SH vaccine formulation is recommended in this whole area. Spatial demes (22 sub-locations) in Southeastern Asia are presented at the bottom.

**Extended Data Fig 2.**
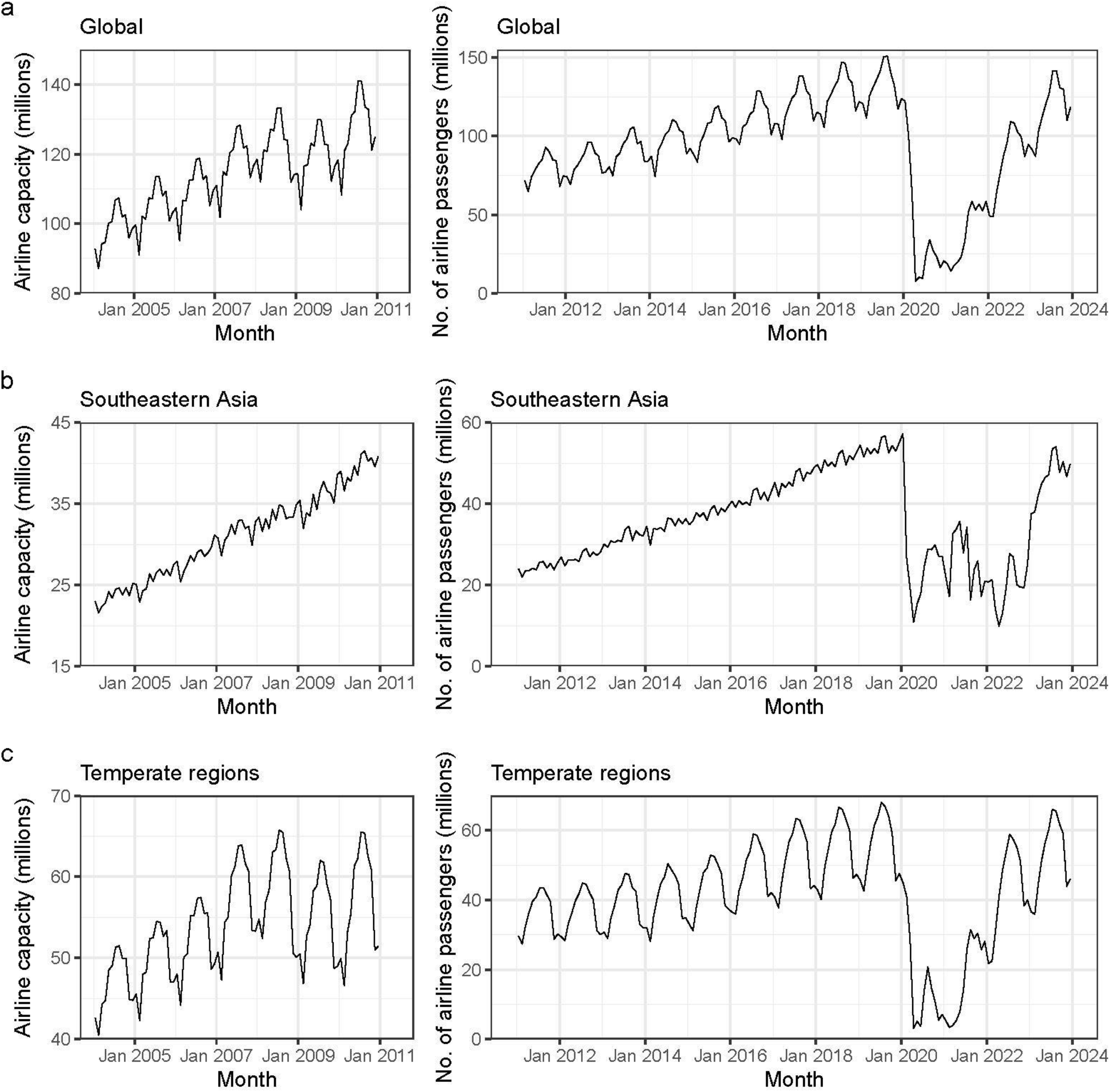
Airline capacity or passengers over time. **a)** Total global monthly airline capacity or passengers. Data of airline passengers are not available before 2011, therefore airline capacity data (number of available seats) are alternatively used; **b)** Total monthly airline capacity or passengers among 22 sub-locations within Southeastern Asia; **c)** Total monthly airline capacity or passengers among countries within temperate regions.

**Extended Data Fig 3.**
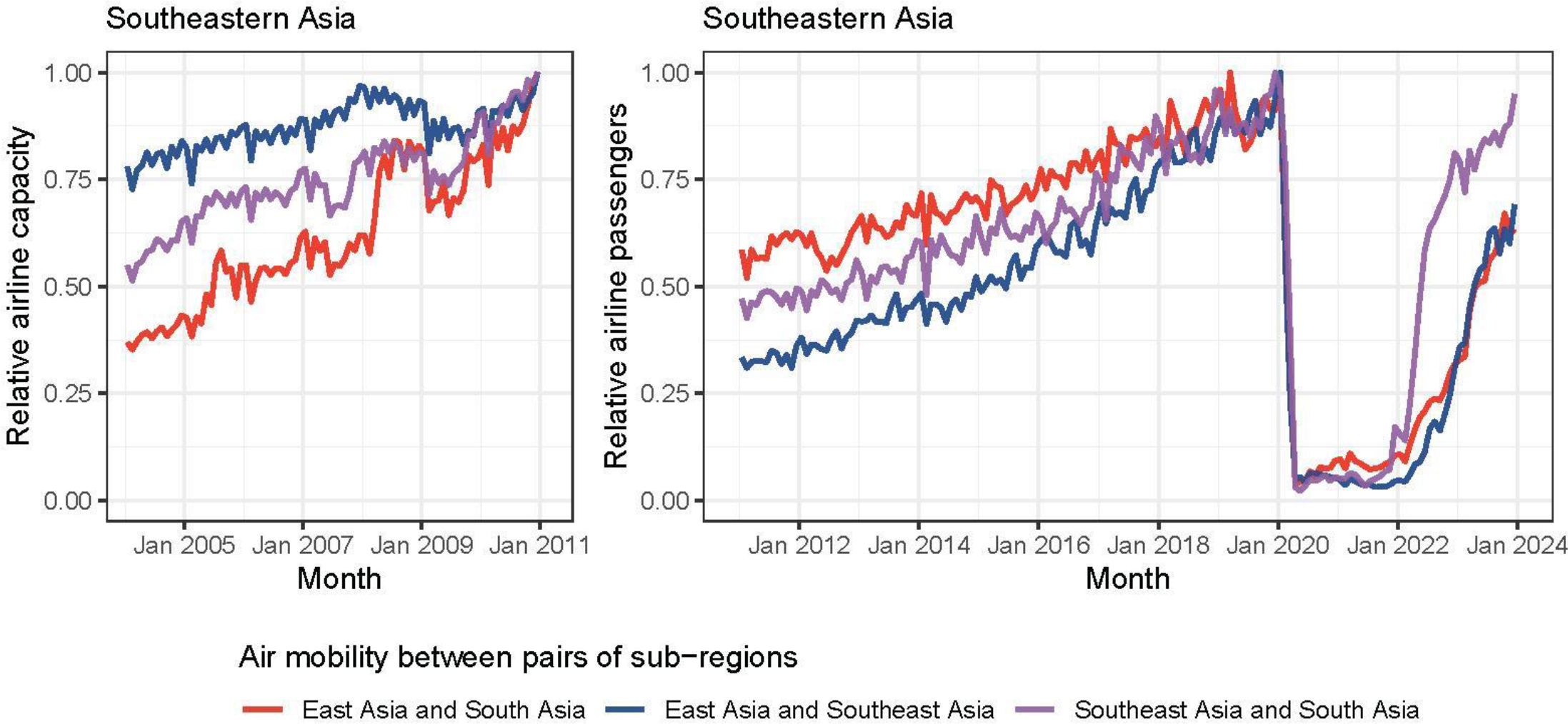
Relative air mobility between pairs of sub-regions with Southeastern Asia. Data of airline passengers are not available before 2011, therefore airline capacity data (number of available seats) are alternatively used. For comparison, each line was divided by its maximum value during the period to obtain the relative value.

**Extended Data Fig 4.**
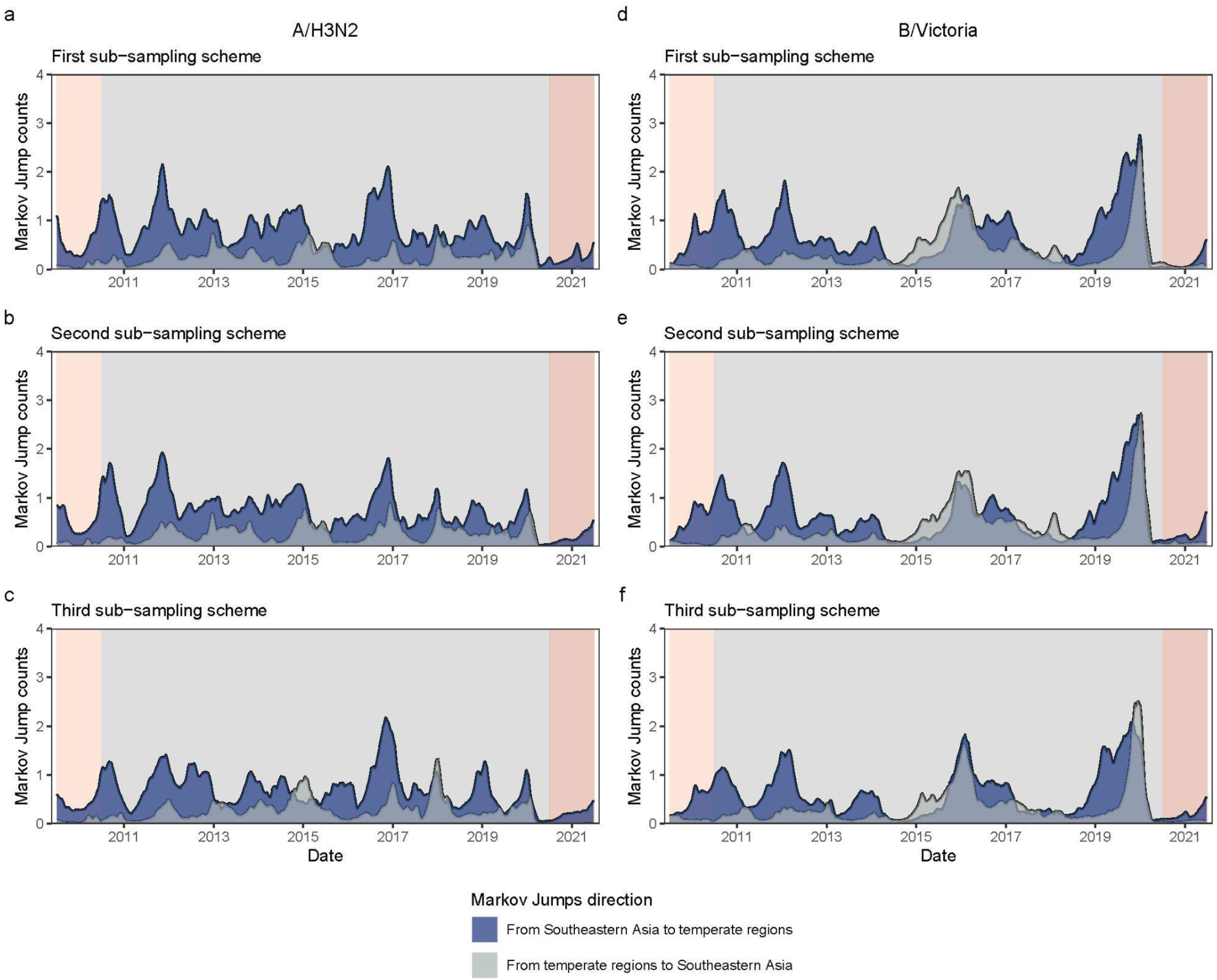
A five-week running average of the weekly Markov Jump counts of A/H3N2 and B/Victoria between two regions using various sets of genetic data from three sub-sampling schemes. Light and dark red shading denote the A/H1N1 and COVID-19 pandemic seasons, respectively, between which are interpandemic seasons. Three sub-sampling schemes refer to even sampling strategy, strategy accounting for population size, and strategy accounting for the product of population size and influenza positivity rate, respectively.

**Extended Data Fig 5.**
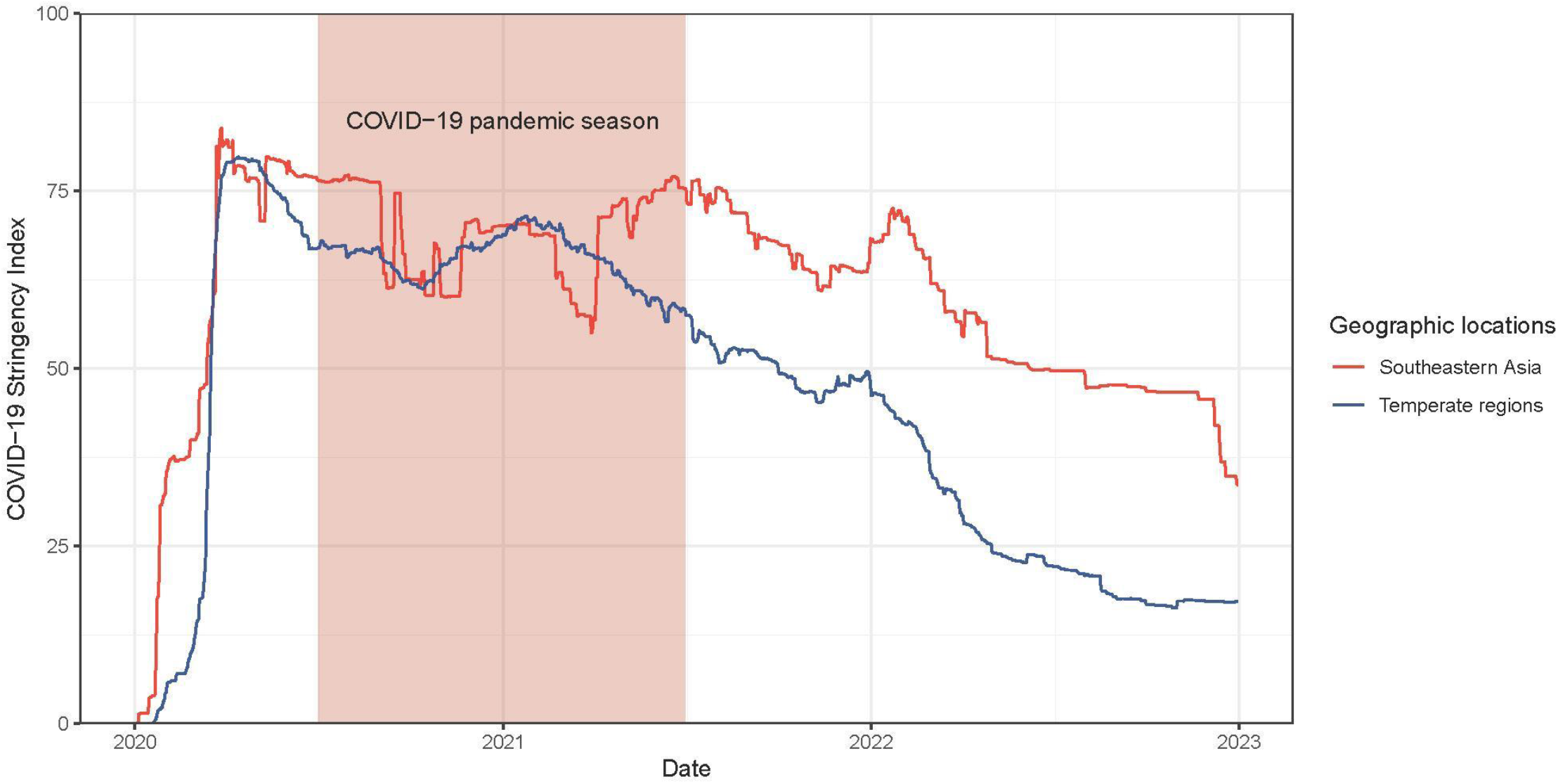
The COVID-19 Stringency Index weighted by population size in Southeastern Asia and temperate regions in the context of COVID-19 pandemic. The data was retrieved from Our World in Data^61^.

**Extended Data Fig 6.**
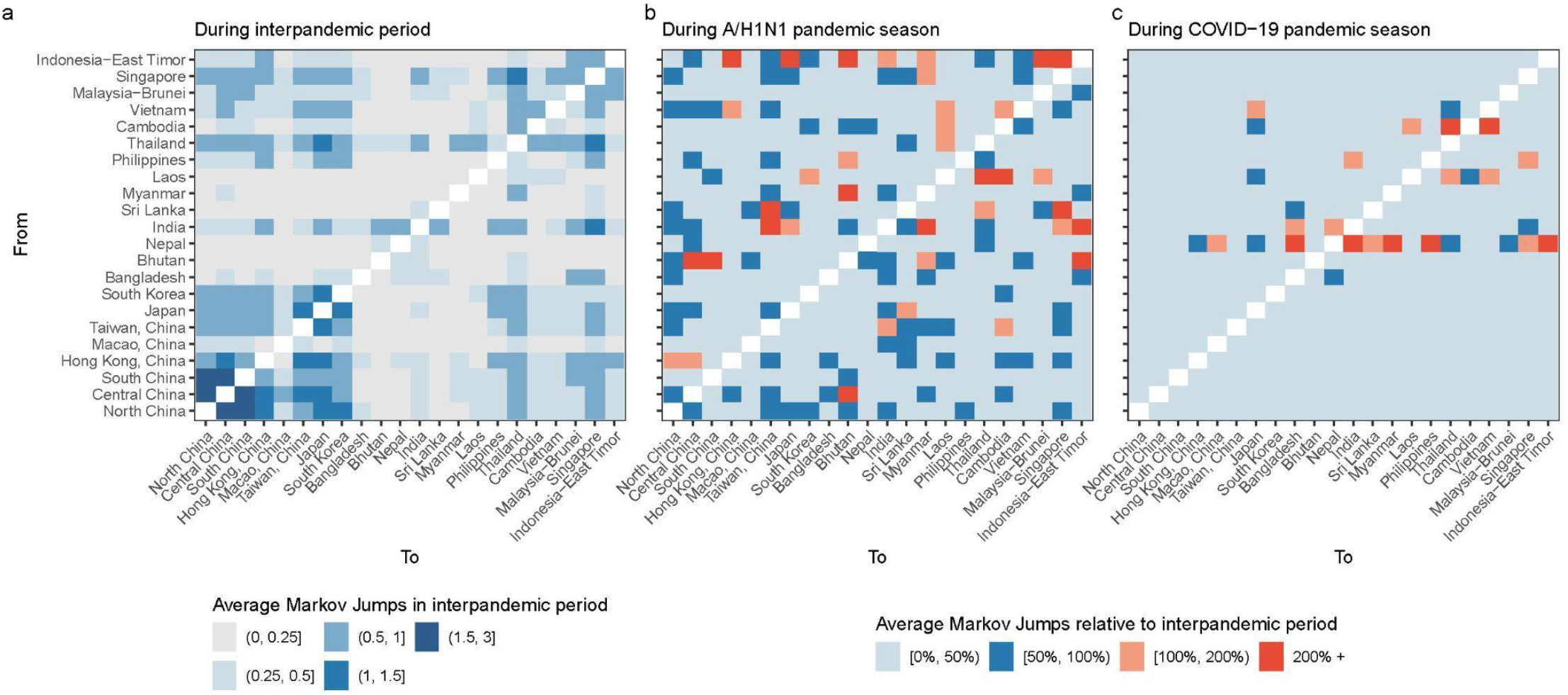
Between-location viral movements of A/H3N2 persistent lineages circulating within Southeastern Asia. **a)** Count of average Markov jumps per season in the interpandemic period; **b-c)** Changes in average Markov jumps during the A/H1N1 and COVID-19 pandemic seasons relative to the interpandemic period.

**Extended Data Fig 7.**
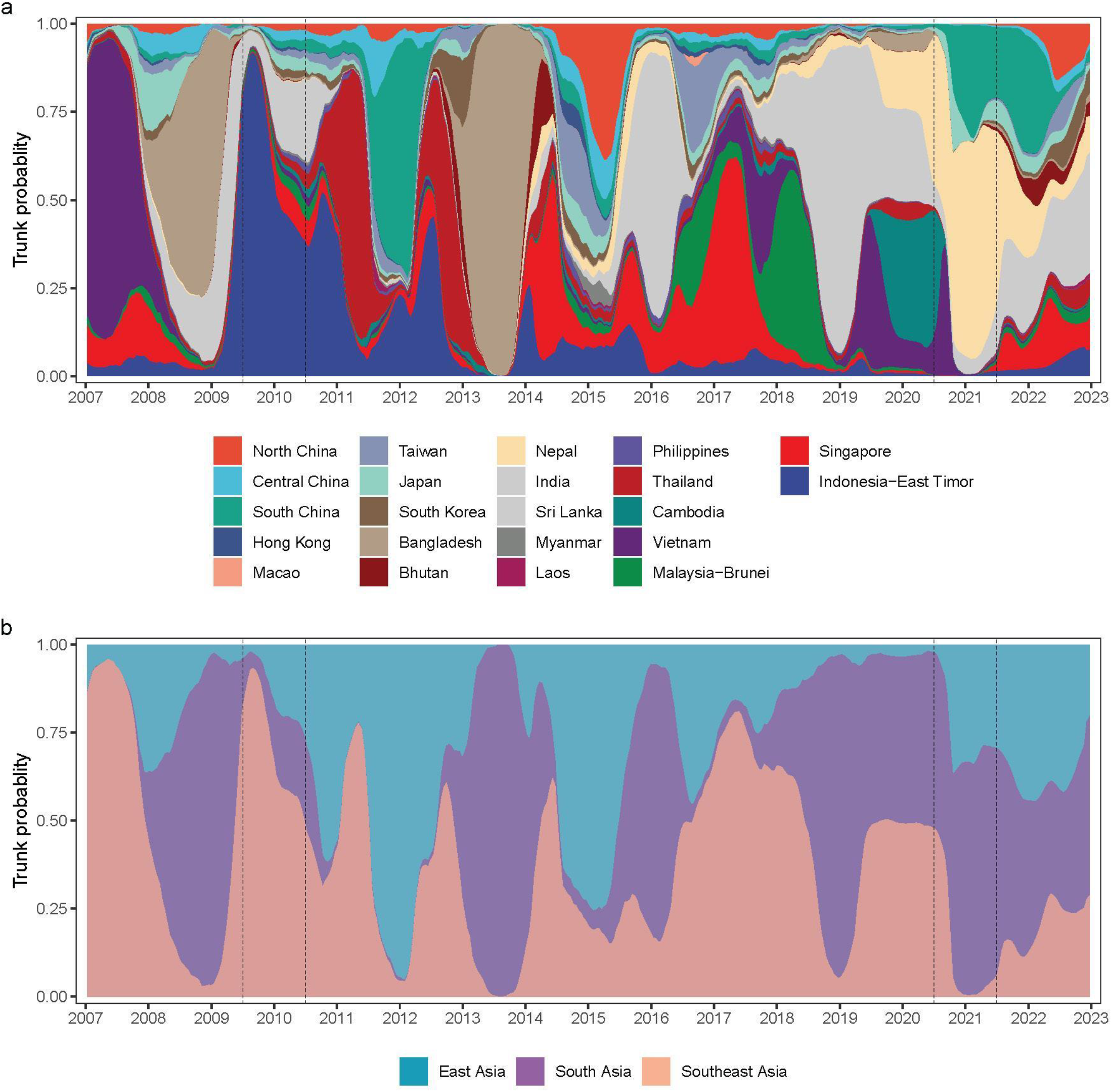
The geographic trunk probability of A/H3N2 persistent lineages circulating in Southeastern Asia. **a)** Trunk location inferred at 22 sub-location levels. **b)** Trunk location aggregated at 3 sub-region levels. Trunk was defined as all branches ancestral to viruses sampled within 1 year of the most recent sample. Vertical dashed black lines refer to the A/H1N1 pandemic and COVID-19 pandemic season, respectively

**Extended Data Fig 8.**
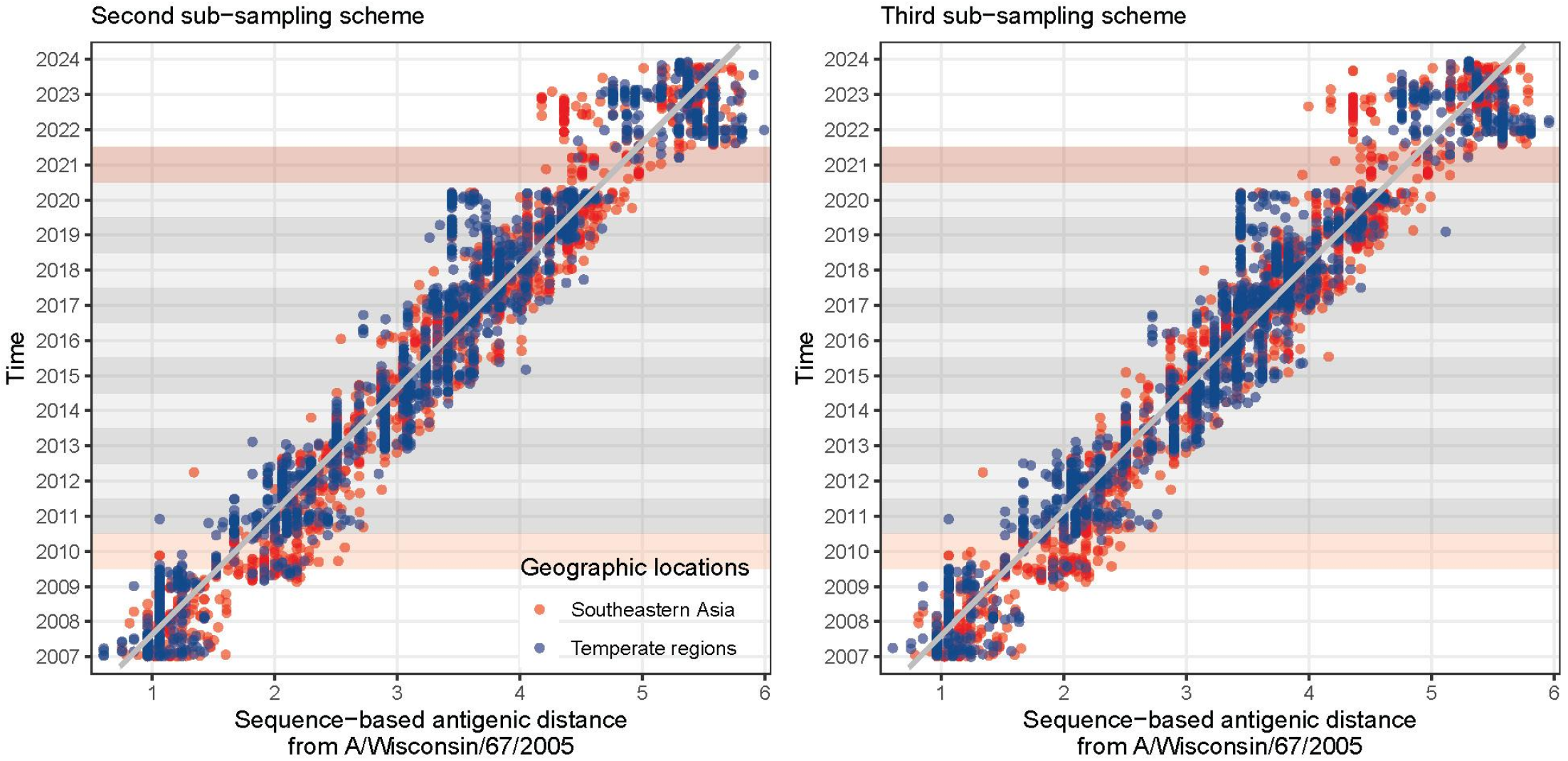
Sequence-based antigenic distance from A/Wisconsin/67/2005 of all virus strains plotted against time of collection using various sets of genetic data from other two sub-sampling schemes. The thick grey line is the fitted line using a linear model. Points to the right of the line are antigenically advanced, whereas strains to the left of the line are antigenically lagging. Light and dark red shading denotes the A/H1N1 and COVID-19 pandemic seasons, respectively, between which were interpandemic seasons.

**Extended Data Table 1.** Statistical test of antigenic distance of A/H3N2 to the fitted line during each pandemic season against interpandemic seasons.

| Influenza seasons | Southeastern Asia |  |  | Temperate regions |  |  |
| --- | --- | --- | --- | --- | --- | --- |
|  | Distance (Months)* | *p* value |  | Distance (Months) | *p* value |  |
| Pandemic seasons |  |  |  |  |  |  |
| 2009/2010 | 0.258 (10.8) | Reference | - | 0.159 (6.7) | Reference | - |
| 2020/2021 | -0.105 (-4.4) | - | Reference | -0.096 (-4.1) | - | Reference |
| Interpandemic seasons | 0.020 (0.8) | < 0.001<sup>‡</sup> | < 0.001 | -0.054 (-2.3) | < 0.001 | 0.632 |

\*Average antigenic distance of A/H3N2 virus lineages circulating in specific season and region to the fitted line (Fig. 4a), for which the line can also be interpreted as a function of time (months). Positive values refer to antigenically advanced distance (months), whereas negative values refer to antigenically lagging distance (months).
<sup>‡</sup> It is the comparison of antigenic distance of virus lineages circulating between pandemic season (2009/2010, as reference) and all interpandemic seasons, where a *p* value was yielded by Bootstrap resampling of 1,000 samples.

## Supplementary information

**Supplementary Fig 1.**
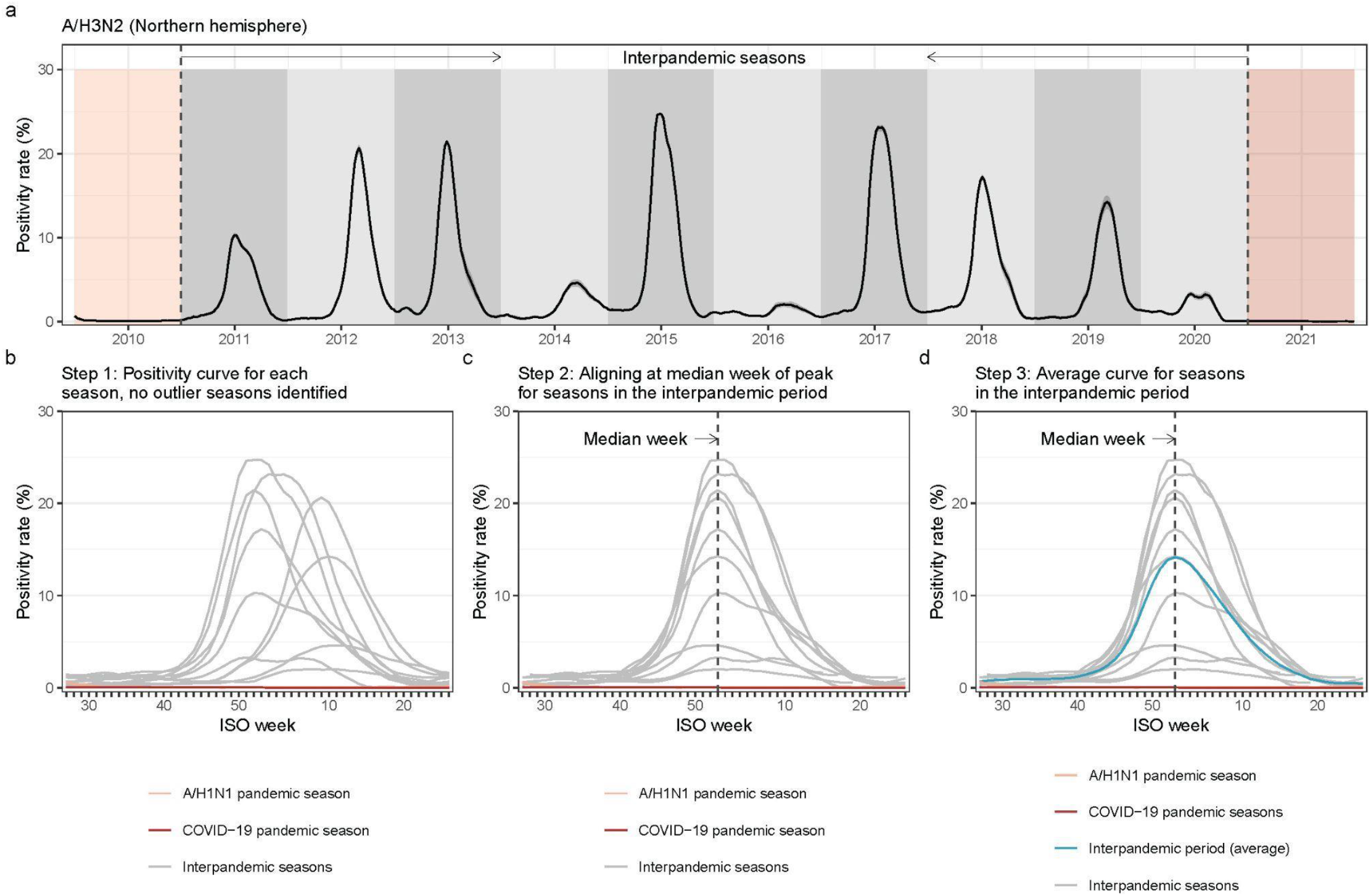
Illustration of the methodology used to determine the average curve of A/H3N2 positivity rate in the Northern hemisphere for seasons in the interpandemic period. **a**) A/H3N2 positivity rates over time in the Northern hemisphere. Light and dark red shading denotes the A/H1N1 and COVID-19 pandemic seasons, respectively. **b**) Step 1: Plotting the positivity curve for each season, where the season was defined as running from ISO week 27 of one year to ISO week 26 of the next year. No outlier seasons (defined as a season where the peak positivity rate occurred at the start or end of the season, i.e., ISO week 26 or 27 for the Northern hemisphere) were identified during the interpandemic period. **c**) Step 2: Identifying the median week of peak occurrence (ISO week 1, here) for the seasons in the interpandemic period after removing the outlier seasons, followed by the epidemic alignment of interpandemic seasons. **d**) Step 3: Calculating an average curve for seasons in the interpandemic period.

**Supplementary Fig 2.**
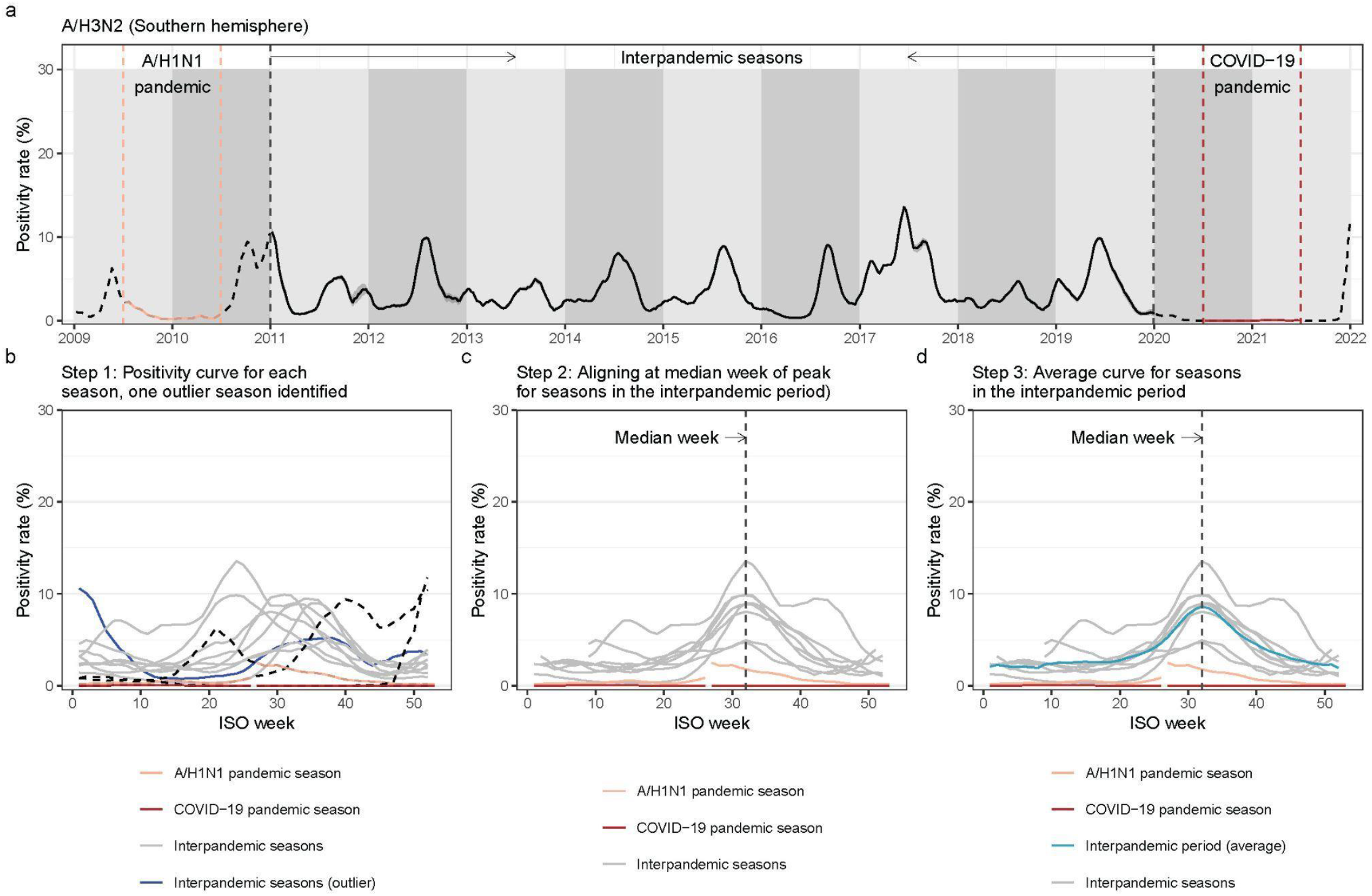
Illustration of the methodology used to determine the average curve of A/H3N2 positivity rate in the Southern hemisphere for seasons in the interpandemic period. **a**) A/H3N2 positivity rates over time in the Southern hemisphere (SH). Light and dark red lines denote the A/H1N1 and COVID-19 pandemic seasons, respectively. **b**) Step 1: Plotting the positivity curve for each season, where the season was defined as running from ISO week 1 to week 52 of one year. One outlier season (dark blue line, corresponding to the 2011 season) was identified since the peak positivity rate occurred at the start of the season. The pandemic seasons defined here both spanned across two SH seasons, so dotted lines (Jan-Jun 2009, Jul-Dec 2010, Jan-Jun 2020, Jul-Dec 2021) are used to refer to curves outside the pandemic influenza seasons. **c**) Step 2: Identifying the median week of peak occurrence (ISO week 32, here) for the seasons in the interpandemic period, after removing the outlier season, followed by the epidemic alignment of interpandemic seasons. **d**) Step 3: Calculating an average curve for seasons in the interpandemic period. The first half of the positivity lines (Jan-Jun 2010; Jan-Jun 2021) during the pandemic periods occurred after the second half of the line (Jul-Dec 2009; Jul-Dec 2020).

